# Estimates of the Prevalence of Autism Spectrum Disorder in the Middle East and North Africa Region: A Systematic Review and Meta-Analysis

**DOI:** 10.1101/2024.08.27.24312604

**Authors:** Aishat F. Akomolafe, Bushra M. Abdallah, Fathima R. Mahmood, Amgad M. Elshoeibi, Aisha Abdulla Al-Khulaifi, Elhassan Mahmoud, Yara Dweidri, Nour Darwish, Duaa Yousif, Hafsa Khalid, Majed Al-Theyab, Muhammad Waqar Azeem, Durre Shahwar, Madeeha Kamal, Majid Alabdulla, Salma M. Khaled, Tawanda Chivese

**Author notes:** Corresponding and guarantor author: Tawanda Chivese, Department of Population Medicine, College of Medicine, QU Health, Qatar University, Doha, Qatar, P O BOX 2713, Doha, Qatar. These authors contributed equally (Aishat F. Akomolafe, Bushra M. Abdallah, Fathima R. Mahmood).

## Abstract

**Background and Objective:** Estimates of the prevalence of the autism spectrum disorder (ASD) in the Middle East and North Africa (MENA) region are not readily available, amid a lack of recent evidence. In this study, we estimated the prevalence of ASD in the MENA region by synthesising evidence from published studies in the region.

**Methods:** In this systematic review and meta-analysis, we searched PubMed, EMBASE, Scopus, and CINAHL databases for studies which assessed ASD prevalence in the MENA region. Risk of bias was assessed using the Newcastle Ottawa scale. A bias-adjusted inverse variance heterogeneity meta-analysis model was used to pool prevalence estimates from included studies. Cochran’s Q statistic and the I^2^ statistic were used to assess heterogeneity, and publication bias assessed using funnel and Doi plots.

**Results:** A total of 3075 studies were identified, 16 studies of which met the inclusion criteria and involved 3,727,731 individuals. The studies were published during the period 2007-2022, and included individuals from Iran, Oman, Libya, Egypt, Kingdom of Saudi Arabia (KSA), Lebanon, United Arab Emirates (UAE), Bahrain and Qatar. Estimates of ASD prevalence ranged from 0.01% in Oman during the period June 2009-December 2009, to a high of 2.51% in the Kingdom of Saudi Arabia during the period December 2017-March 2018. The pooled prevalence of ASD was 0.13% (95% CI: 0.01% – 0.33%), with significant heterogeneity (I^2^ = 99.8%). For Iran, the only country with multiple analysable studies, an overall prevalence of 0.06% (95% CI: 0.00 – 0.19, I2=97.5%, n= 6 studies) was found. A review of data from countries with repeated studies suggested that the prevalence of ASD is increasing.

**Conclusion:** Estimates of the prevalence of ASD vary widely across the MENA region, from 0.01% in Oman to 2.51% in Saudi, with an overall prevalence of 0.13%. Existing data suggests a trend towards increasing prevalence in the region. More and better-quality research is needed to provide up to date ASD prevalence estimates.

**Registration:** The protocol for this systematic review and meta-analysis was registered on the International Prospective Register of Systematic Reviews (PROSPERO) with registration ID CRD42024499837.

## 1. Introduction

The autism spectrum disorder (ASD) is a neurological and developmental disorder whose main features include difficulties with social communication and social interactions, restricted, repetitive behavioural patterns, and limited interests (1). These symptoms usually manifest in early childhood and persist throughout life. ASD carries significant financial, social, and health-related burden to individuals, families, and society. Families of individuals with ASD often face significant emotional and financial challenges due to high levels of anxiety, stress, and isolation associated with caregiving, as well as the extensive financial resources required (2). Co-occurring conditions, such as anxiety disorders, attention deficit/hyperactivity disorder (ADHD), depression, intellectual disability and seizures are common among individuals with ASD (3), with one study showing that over 70% of individuals with ASD have at least one co-occurring psychiatric condition, and more than 40% have two or more (4).

ASD affects all regional, racial, and socioeconomic groups. The World Health Organization (WHO) estimates the global prevalence of ASD in 2023 to be approximately 1 in 100 children worldwide (5). However, ASD prevalence rates vary globally due to several reasons, which include variation in awareness in different countries, the absence of diagnostic tools that are culture sensitive, and cultural variation in interpreting the behaviour of children (2, 6). The reported prevalence of ASD in the MENA region tends to be lower than in Western countries (7). This could be because of underestimation of ASD prevalence due to social stigma, limited access to diagnostic resources, and poor screening programs in some MENA countries (1).

Recent epidemiological studies indicate a significant global increase in the annual prevalence of ASD over the past few years (8). According to a 2023 systemic analysis investigating the global burden of ASD, the prevalence of autism in the MENA region has witnessed a 70% increase between 1990 to 2019, comprising 7.5% of the new ASD cases globally (1). This increase has been attributed to several reasons including an expanded definition of the spectrum, updates in diagnostic criteria and screening tools, shifts in research methodologies, and greater awareness of ASD (4). Despite this noted increase in ASD prevalence, studies on ASD prevalence in the MENA region are still limited, and current estimates of ASD prevalence are lacking. Even the aforementioned systemic analysis (1) left out many existing prevalence studies from the Middle East and North Africa region. This highlights the need for an up-to-date systematic review and meta-analyses to estimate the prevalence of ASD in the MENA region, which is the aim of the present study.

## 2. Methods

### 2.1. Study Design and Protocol Registration

A systematic review and meta-analysis of studies that met the eligibility criteria was carried out. The study adheres to the Preferred Reporting Items for Systematic Reviews and Meta-Analyses (Supplementary Table S1) (9). The study protocol is registered in the International Prospective Register of Systematic Reviews (PROSPERO) with the registration number CRD42024499837.

### 2.2. Search Strategy and Study Selection

We searched PubMed, EMBASE, Scopus, and Cumulative Index to Nursing and Allied Health Literature (CINAHL) for relevant articles published until 20 January 2024. We used medical subject headings or MeSH terms and keyword searches for ASD and the Middle East and North Africa (MENA) region in the PubMed search. No language restrictions were imposed. The detailed search strategy is shown in Supplementary Tables S2 – S5. After removing duplicates using EndNote, Rayyan systematic review management website (www.rayyan.ai) was used to screen studies for inclusion. Within Rayyan, two reviewers independently screened each study for inclusion based on title and abstract. After initial screening, two reviewers independently screened the full text of each study for inclusion, based on the specified inclusion criteria. In cases of disagreement between reviewers, a third reviewer was consulted.

### 2.3. Eligibility Criteria

This systematic review and meta-analysis included observational studies that investigated the prevalence of ASD in the MENA region. Studies were included if they documented the prevalence of ASD. Studies were excluded if the study population included participants living outside of the MENA region. Studies were also excluded if they were case control studies, case series, case reports, letters, opinions, narrative reviews, or other studies that did not contain primary data, if they were duplicates, or if they did not have full text. Exceptions were two case control studies which estimated prevalence by dividing the number of cases (100) by the estimated population for that year.

### 2.4. Data Extraction

In case of duplicate publications, the article that contained the most information was included in the review and all others were excluded as duplicates. The data extracted from the studies included: publication year, country, study design, study period, ASD screening tool used, mean age of participants, and sample size. The prevalence was extracted as the number of ASD cases (numerator) over the total sample size (denominator). For each study, two reviewers independently extracted the data into a standardized data extraction sheet using Microsoft Office Excel. Disparities in data extracted were resolved via discussion between the reviewers.

### 2.5. Assessment of the Quality of Included Studies

The quality of the included studies was assessed using a modified version of the Newcastle Ottawa Scale (NOS) (Supplementary Tables S6 – S7). The NOS assesses selection, comparability, and exposure/outcome, with scores ranging from 0 to 9 points (10). The modified version of the scale for cross sectional studies had scores ranging from 0 to 15 points. Studies were categorized based on quality scores, with scores between 13 to 15 indicating high quality, 8 to 12 indicating moderate quality, and scores below 7 indicating low quality. Each included study was independently evaluated by two reviewers. In case of discrepancies, a third reviewer was consulted.

### 2.6. Synthesis of Findings

We narratively described qualitative data, including author names, year of publication, country, study design, and sample size in tables. Unadjusted prevalence estimates of ASD and their 95% confidence intervals were calculated for each included study, and they were then pooled, when possible. Where overall synthesis was not possible, a systematic review of the prevalence estimates was done. The quality-effects model was used for the subsequent meta-analysis (11). This model depended on the use of additional data, including the quality ranking of each study, to modify the variance weights. The modified NOS was used to calculate the quality weights. In the meta-analysis, the variance of the prevalence data was stabilized using the Freeman-Tukey transformation. To assess the robustness of the results, sensitivity analysis using leave one out analyses was also performed. Forest plots were used to show the pooled prevalence estimate. Cochran’s Q p-values and the I^2^ statistic were used to measure heterogeneity (12). I^2^ values of 25%, 50%, and 75%, respectively, were taken to represent low, moderate, and high levels of statistical inconsistency. Funnel and Doi plots were used to evaluate publication bias. The LFK index was used to assess symmetry in Doi plots, with a value greater than 1 or less than -1 indicating minor asymmetry, and a value greater than 2 or less than -2 indicating major asymmetry. (13). Exact p-values were reported. Stata statistical software was used to conduct all analyses, and PRISMA guidelines (14) were followed.

## 3. Results

### 3.1. Search Results

A total of 3076 study reports were identified from database searches. Out of these, 219 studies remained after excluding duplicates, irrelevant studies, and those which the full text could not be retrieved (Fig.1, and Supplementary Table S8). The final number of studies included in the systemic review was 16 (15–30).

**Fig. 1.**
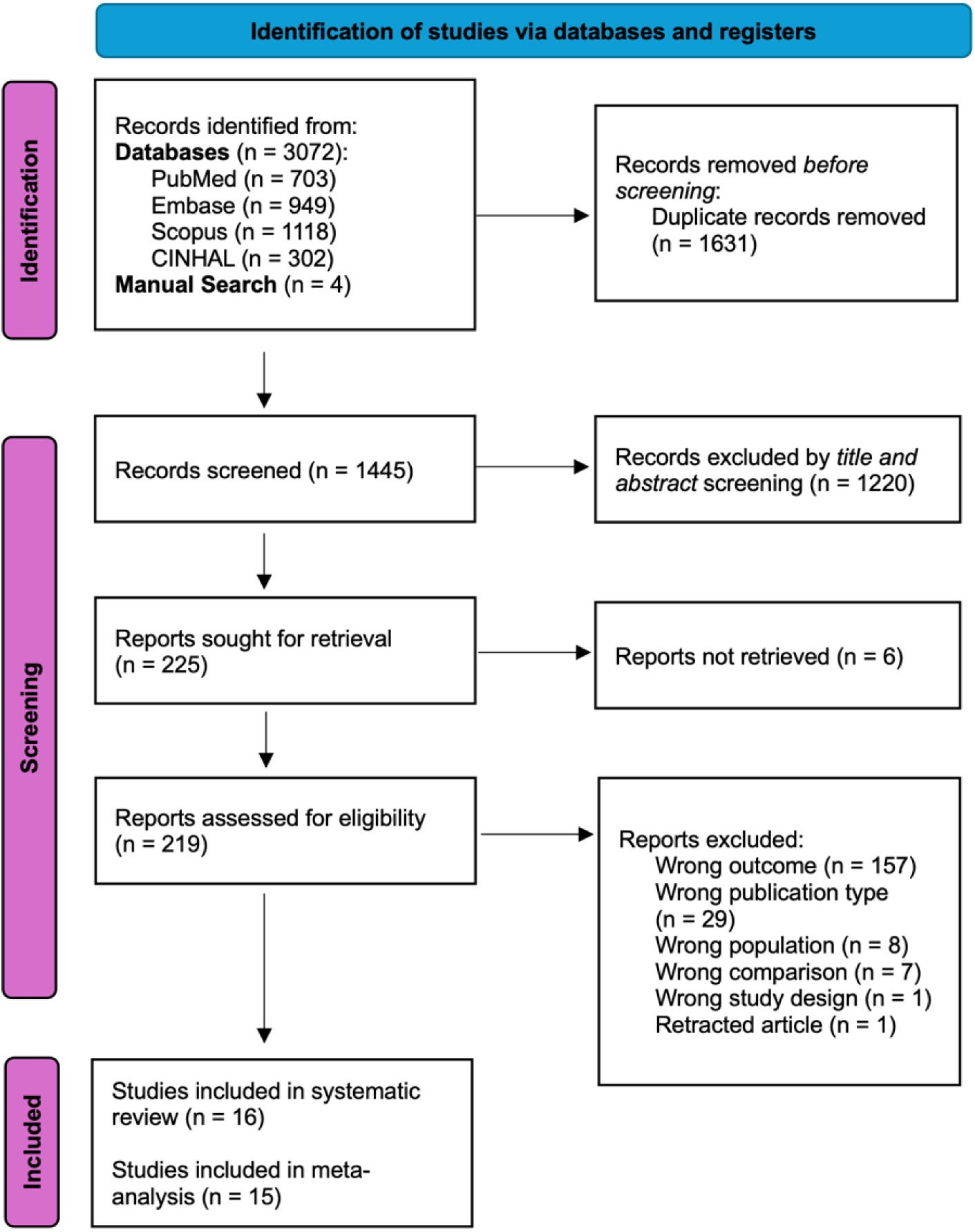
PRISMA flowchart.

### 3.2. Characteristics of Included Studies

The characteristics of the 16 studies in the systematic review are shown in Table 1. Most of the studies were from Iran (n=6) (17, 21–23, 25, 26), two were from Oman (15, 18) and two from the Kingdom of Saudi Arabia (24, 30). The following countries contributed one study each to the analysis: Qatar (19), Egypt (27), Lebanon (20), Bahrain (29), United Arab Emirates (16), and Libya (28). Most of the studies were published during the year 2019 (n=5). There was one study from 2007, 2010, 2011, 2013, 2015, 2016, 2022, and 2 from 2012 and 2021.

**Table 1:** Characteristics of Included Studies.

| Study | Prevalence Year | Country | Study Design | Total Number of Participants | ASD Number of participants | Mean age (SD) of participants (in years) | % of males in study | Screening tool used | ASD prevalence (per 10,000) |
| --- | --- | --- | --- | --- | --- | --- | --- | --- | --- |
| Eapen, 2007 (16) | Not specified | UAE | cross sectional | 694 | 3 | 3 years and 7 months | 49.4 | DSM-IV | 29 |
| Sasanfar, 2010 (17) | 2005 | Iran | case-cohort | 200000 | 219 | Age range 5-11 years (mean age not specified) | 51.6 | SCQ and DSM-IV TR | 11 |
| Al Farsi, 2011 (15) | 2009 | Oman | cross sectional | 798,913 | 113 | Age range 0–14 years | 74.3 | DSM-IV-TR | 1.4 |
| Samadi, 2012 (25) | 2006-2009 | Iran | cross sectional | 1,320,334 | 826 | 5-year-olds | 51.4 | ADI-R | 6.3 |
| Zeglam, 2012 (28) | 2005-2009 | Libya | cross sectional | 38 508 | 128 | median age was 4 years and 6 months | Not stated | DSM-IV | 30 |
| Al-Ansari, 2013 (29) | 2005 | Bahrain | Case- control | Not specified | 100 | 10.5 ($\pm 6.4$) (for cases) | 80 (cases only) | M-CHAT, DSM-IV-TR and CARS | 4.3 |
| Samadi, 2015 (26) | 2010 | Iran | cross sectional | 2941 | 28 | 3.3 | 54.8 | ADOS, ADI-R | 95.2 |
| Chaaya, 2016 (20) | 2014 | Lebanon | cross sectional | 998 | 15 | 27.6 ($\pm 6.8$) months | 54 | M-CHAT and a short questionnaire | 153 |
| Goodarzi, 2019 (21) | 2015-2016 | Iran | cross sectional | 1500 | 23 | 25.0 ± 5.3 months (for cases) | 46.9 | M-CHAT | 150 |
| Al- Mamri, 2019 (18) | 2011-2018 | Oman | retrospective descriptive study | 837,655 | 1705 | 4.8 ± 2.4 years (for cases) | 78.1 (cases only) | DSM-5 | 20.4 |
| Mohammadi, 2019 (23) | Not specified | Iran | cross sectional | 31000 | 37 | 6-18 years old | 49 | K-SADS-PL | 10 |
| Manzouri, 2019 (22) | 2017 | Iran | cross sectional | 1504 | 12 | 20.3±3.7 (16-30) in months | 50.2 | M-CHAT-R/F | 80 |
| Alshaban, 2019 (19) | 2015 | Qatar | cross sectional | 133781 | 1525 | Not specified | Not specified | DSM-5 | 114 |
| Sabbagh, 2021(24) | 2020 | KSA | cross sectional | 347,036 | 1023 | 9.0 (2.4) | 76.6 (cases only) | DSM-5 | 28.1 |
| Yousef, 2021 (27) | 2017 | Egypt | cross sectional | 3722 | 2 | Not specified | 50.7 | DSM-5, CARS | 54 |
| Albatti, 2022(30) | 2017-2018 | KSA | cross sectional | 398 | 10 | Age range 2–4 years | 51.7 | M-CHAT-R, ADOS-2 | 250 |

***SCQ- Social Communication Questionnaire; M-CHAT- Modified Checklist for Autism in Toddlers; CARS- Childhood Autism Rating Scale; DSM-IV-TR- Diagnostic and statistical manual of mental disorders, 4th ed, text revision; ADOS-Autism Diagnostic Observation Schedule; ADI- Autism diagnostic interview; K-SADS-PL-The Schedule for Affective Disorders and Schizophrenia for School-Age Children-Present and Lifetime version***

A total of thirteen studies used a cross-sectional design, one study used a case control design, one study used a case-cohort design, and the last study employed a retrospective descriptive design. Case control studies were included only if they calculated prevalence based on their cases and used the country’s population as the denominator rather than cases divided by controls. The case control study (29), was only described in the systematic review and not in the meta-analysis as it did not exclusively state the denominator used in the prevalence calculation. There was variability in diagnostic tools used across studies, with The Diagnostic and Statistical Manual of Mental Disorders, Fourth Edition (DSM-IV) and Modified Checklist for Autism in Toddlers (M-CHAT) being used by five studies each. Other tools used in defining ASD included The Diagnostic and Statistical Manual of Mental Disorders, Fifth Edition (DSM-5), Autism Diagnostic Interview-Revised (ADI-R), Social Communication Questionnaire (SCQ), Autism Diagnostic Observation Schedule (ADOS), Autism Diagnostic Observation Schedule, Second Edition (ADOS-2) and Kiddie Schedule for Affective Disorders and Schizophrenia for School-Age Children-Present and Lifetime Version (2009) (K-SADS-PL-2009).

### 3.3. Assessment of the quality of included studies

The quality assessment of the included studies was assessed using a modified version of the Newcastle-Ottawa Scale (NOS). A total 11 of the included studies were of moderate quality, the scores ranged from 8-15 and the average was 10 out of 15. Most of the cross-sectional studies had safeguards present in 5 of the 8 domains that the NOS scale assessed including representatives of the sample, sample size, ascertainment of the exposure (risk factor), assessment of outcome, and statistical test. While non-respondents, age and sex, and other variables had deficiencies in safeguards. When looking at the case-control studies, overall, both studies had safeguards in representativeness of cases, definition of controls, ascertainment of exposure, and same ascertainment method. The individual quality assessments of all the included studies are shown in Supplementary Tables S9 – S10.

### 3.4. Prevalence of Autism Spectrum Disorder

The metanalysis included a total of 3,717,731 participants, 5687 of whom had ASD. As shown in Figure 2, the overall synthesised prevalence was 0.13% (95% CI 0.01% to 0.33%, n=15 studies), with significant heterogeneity (I^2^=99.8%). Assessment of publication bias showed major asymmetry (Supplementary Figures S1 – S2). On leave-one-out analysis, the prevalence ranged from 0.11%-0.18% after successive removal of each study.

**Fig. 2.**
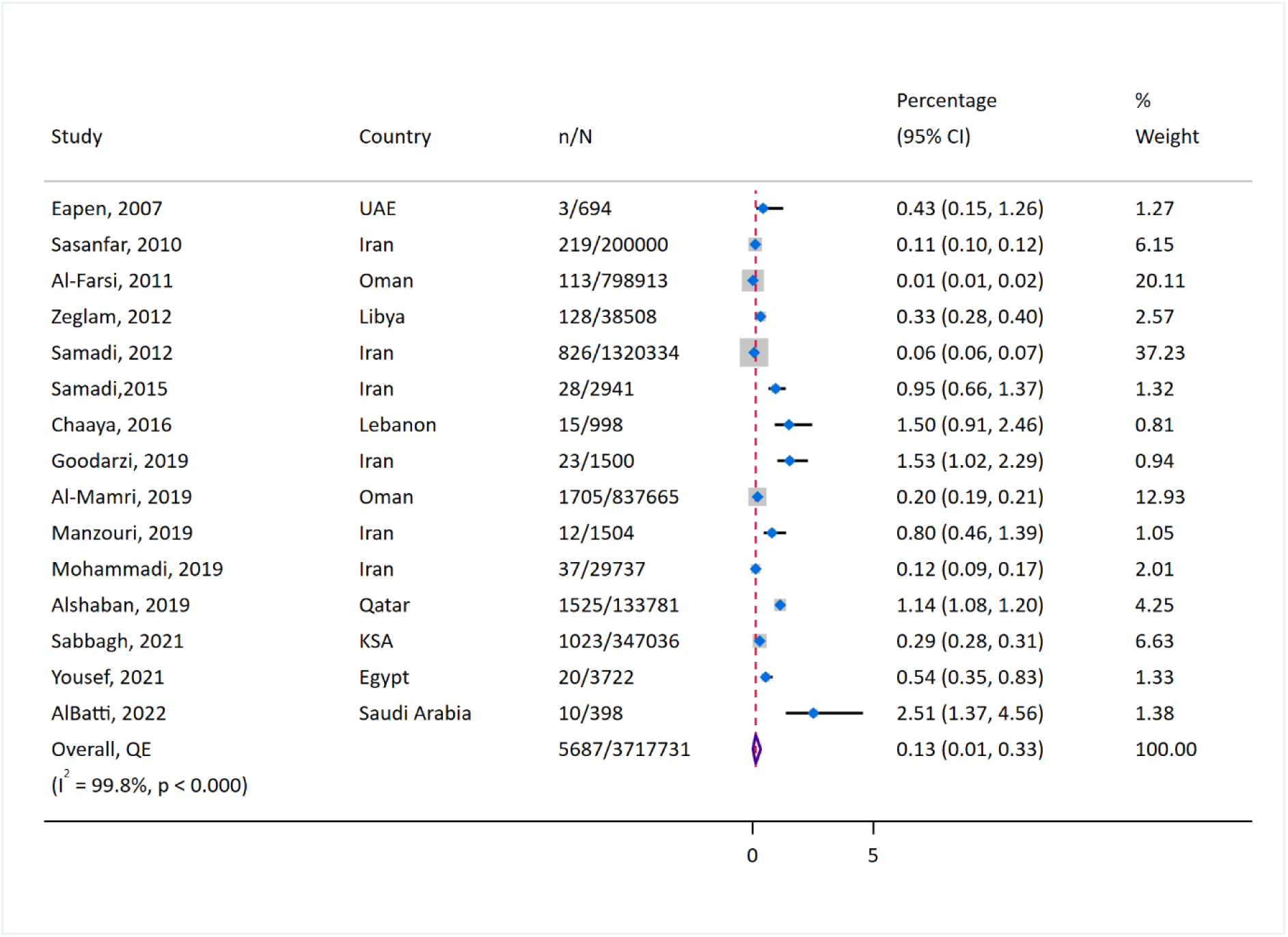
Forest plot of prevalence.

Due to the high heterogeneity observed, a descriptive analysis of ASD prevalence in each country was carried out. The ASD prevalence ranged from 0.01% in Oman to 2.51% in Saudi Arabia. In Qatar, the reported 2015 prevalence was 1.14% (95% CI 1.08%-1.20%)(19). In the UAE, the prevalence of ASD was 0.43% (95% CI 0.15%-1.26%), again from a single study published in 2007 (16). In Oman, the prevalence varied from 0.01% (95% CI 0.01%-0.02%) in 2009 (15) to 0.20% (95% CI 0.19%-0.21%) between the years 2011-2018 (18). Between the years 2005-2009 in Libya, the prevalence was 0.33% (95% CI 0.28%-0.40%) (28). In 2005, in Bahrain, the prevalence was 0.043% (29). In Lebanon, the prevalence was 1.50% (95% CI 0.91%-2.46%) in 2014 (20). In Egypt, the prevalence of ASD was 0.54% (95% CI 0.35%-0.83%) in 2017 (27). In KSA, the prevalence of ASD varied from 2.51% (95% CI 1.37%-4.56%) between the years 2017-2018 (30) to 0.29% (95% CI 0.28%-0.31%) during 2020 (24).

Figure 2 shows that there was a general trend of increasing prevalence of autism in the MENA region with time. Using the year 2015 as a midpoint, the majority of the studies (n=5/6) published before or during 2015 reported a prevalence lower than 0.50% while studies after 2015 tended to report a prevalence greater than 0.50% (n=6/10). Additionally, in Oman and Iran, two of the three countries where multiple prevalence studies were conducted, an increase in the prevalence of ASD was seen. In Oman, there was an increase from 0.01% in 2009 (15) to 0.20 in the prevalence studied between 2011 and 2018 (18). In Iran, the estimated prevalence of ASD increased from 0.11% (95% CI 0.10%-0.12%) in 2005 (17), to 0.80% (95% CI 0.46%-1.39%) in 2017 (22).

### 3.5. Subgroup Analysis

To explore the high heterogeneity, subgroup analysis was carried out by country, but because of sparse data from other countries, the analysis was restricted to Iran only, which had six studies (Fig. 3). In Iran, the prevalence of ASD ranged from 0.06% in a study carried out during 2006-2009 in a population of 1,320,334 to a high of 1.53% in a population of 1500 in a study carried out during 2015-2016. The overall synthesised prevalence was 0.06% (95% CI: 0.00 – 0.19), with significant heterogeneity (I^2^=97.5%), and evidence of publication bias / small study effects (Supplementary Figures S4 – S5). A leave-one-out analysis showed that the Samadi study (25) had the most significant influence on the overall prevalence and the prevalence without this study would have been 0.12% (95% CI: 0.00 – 0.45) (Supplementary Figure S6). Subgroup analysis could not be done for the other countries as there were insufficient studies.

**Fig. 3.**
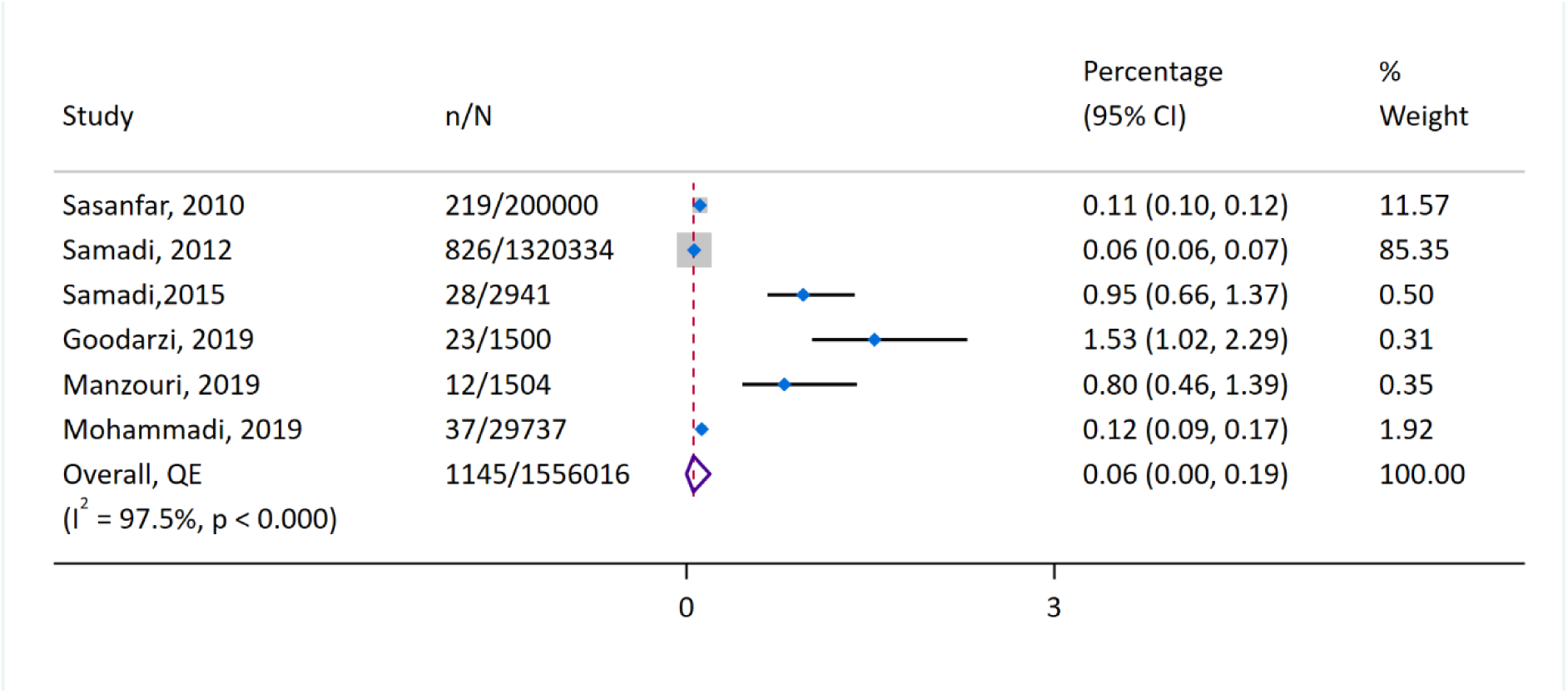
Prevalence in Iran.

### 3.6. Secondary Outcomes

Our research also aimed to examine the severity ratings of ASD among individuals. There were three studies that reported phenotype specific ASD data; one each from Libya, Egypt, and Kingdom of Saudi Arabia. A meta-analysis was not done due to few studies being available. In Libya, out of 180 children referred to Al-Khadra Teaching Hospital for delayed speech and language, 6 of them had Asperger syndrome (28). In Egypt, in a study carried out in the Sharkia Governorate, out of the 104 children at high risk for ASD, 13 were diagnosed with mild to moderate autism and 7 were diagnosed with severe autism (27). In KSA, out of 205 children diagnosed with autism in Makkah and Jeddah, 62 were diagnosed with mild autism, 106 with moderate autism and 37 with severe autism (27).

## 4. Discussion

Our systematic review and meta-analysis of 16 studies and a total of 3,717,731 participants found the prevalence of ASD ranging from 0.01% in Oman to 2.51% in Saudi Arabia and an overall prevalence of 0.13%. Our analysis revealed significantly high heterogeneity which was expected due to differences in prevalence measurement methods and diagnostic tools employed by each study included in the metanalysis.

The overall prevalence of ASD in the MENA region reported in the present study of 0.1% is much lower than the global prevalence calculated by WHO of 1%. Other metanalyses also report a lower estimate of ASD prevalence in the MENA region similarly to our metanalysis. A regional metanalysis conducted in 2014 by Salhia et. al. estimated the prevalence of ASD in the Gulf region to be between 0.014% to 0.29% (31). A more recent 2022 meta-analysis on global ASD prevalence estimated the ASD prevalence to be 1% (95% CI 0.3-3.1%) in Africa, and 0.4% (95% CI 0.1-1%) in Asia with an overall worldwide prevalence of 0.6% (95% CI 0.4-1%) (2). Notably, these existing meta-analyses did not include many of our included studies and therefore the current study offers the most up-to-date estimates of ASD prevalence in the region.

We observed a wide difference in the individual prevalence of MENA countries in our systematic review with prevalence rates ranging 0.01% in Oman to 2.51% in Saudi Arabia. The differences in the individual prevalence could be due to the different screening tools employed and the time periods and populations investigated by each study. Additionally, different sampling methods were used by each study, with studies that employed convenient sampling from paediatric outpatient clinics (21, 30) tending to report higher prevalence estimates than studies that employed random sampling (16, 22, 27).

Some of the between-country prevalence differences could be possibly explained by several reasons. Besides the likelihood of underdiagnosis in low- and middle-income countries due to societal stigma and poor access to healthcare (32), the risk factors for ASD could also differ by country with certain risk factors being more prevalent in specific countries. Several risk factors have been attributed to the pathogenesis of ASD including genetic (33) and parental factors such as advanced maternal age (34), consanguinity (35), and environmental factors such as infections during pregnancy (36), and the distribution of these risk factors vary between countries.

For the only country with more than two studies, Iran, the overall prevalence was 0.06%, again with significant heterogeneity. Although there was a good number of studies reporting on prevalence of ASD in Iran, we could not find an existing metanalysis. However, the 2022 global prevalence systematic review reported a prevalence of 0.063% in Iran, but this was based on only one study (25) which was also included in our meta-analysis (37). The six Iranian studies included in our metanalysis were conducted at different time periods to one another and also differed in the choice of ASD screening tools and the type of prevalence investigated.

With regards to the 70% increase in the prevalence of autism in the MENA region reported by the 2023 global burden study (1), it is important to note that while some studies from our systematic review suggest an increase, more robust evidence is required. A better estimation of the prevalence of ASD is provided by the studies that screened all members of a selected sample used for the denominator to calculate the prevalence compared to those that used the estimated total population. Therefore, future research should focus on screening all members of the sample or selected population to yield higher-quality data.

Our review has several strengths. In this study, we provide the most recent estimates of the prevalence of ASD in the MENA region. Our included 16 studies are the biggest yet in all analyses from the region and the study employs a bias-adjusted synthesis method to weight and rank the studies based on methodological quality. This method is considered more robust than the random effects model when dealing with heterogeneous studies (38). However, there are some limitations. Firstly, a few of the included studies calculated their prevalence as the number of participants with ASD divided by the population of the country at a certain time, which may underestimate the true prevalence. Secondly, we were unable to conduct subgroup analyses by gender or country, with the exception of Iran, due to insufficient data. We also observed a paucity of data regarding the prevalence of ASD in general and of the different subtypes and severity of autism in the MENA region. Therefore, future studies should aim to fill these literature gaps.

## 5. Conclusion

Estimates of the prevalence of ASD varied widely across the MENA region, from 0.01% in Oman to 2.51% in Saudi, with an overall prevalence of 0.13%. Existing data suggests a trend towards increasing prevalence in the region. In the future, more and better-quality research should explore the causes of these observed differences and provide up to date ASD prevalence estimates. Our findings also highlight the variability in ASD prevalence within the MENA region and underscore the need for standardized diagnostic criteria and reporting methods to better understand and address the burden of ASD across different populations in the region.

Further research is warranted to explore the underlying factors contributing to the observed differences and to develop region-specific strategies for early diagnosis and intervention.

## Data Availability

This study is a meta-analysis of published studies and the data used in this work are prevalence rates from the included studies. The availability of the underlying individual participant data is subject to the primary publications' journal policies.

## 6. Ethics

This review used secondary data from peer-reviewed published studies and does not require ethical clearance.

## 9. Author Contributions

Aishat F. Akomolafe: Conceptualization, Methodology, Writing - Original Draft, Writing - Review & Editing, Project administration. Bushra M. Abdallah: Methodology, Writing - Original Draft, Writing - Review & Editing, Project administration. Fathima R. Mahmood: Conceptualization, Methodology, Writing - Original Draft, Writing - Review & Editing, Project administration. Amgad M. Elshoeibi: Methodology, Software, Formal analysis, Data Curation. Elhassan Mahmoud: Methodology, Software, Formal analysis. Aisha Al-Khulaifi: Methodology, Writing - Original Draft. Nour Darwish: Methodology, Writing - Original Draft. Yara Dweidri: Methodology, Writing - Original Draft. Duaa Yousif: Methodology, Writing - Original Draft. Hafsa Khalid: Methodology, Writing - Original Draft. Majed Al-Theyab: Methodology, Writing - Original Draft. Muhammad Waqar Azeem: Writing - Review & Editing. Madeeha Kamal: Writing - Review & Editing. Durre Shahwar: Writing - Review & Editing. Majid Alabdulla: Writing - Review & Editing. Salma M. Khaled: Supervision, Writing - Review & Editing Project administration, Funding acquisition. Tawanda Chivese: Conceptualization, Methodology, Writing - Review & Editing, Formal analysis, Supervision, Project administration, Funding acquisition. All authors made a significant contribution to the work reported and gave final approval of the version to be published.

## References

1. Ebrahimi Meimand S, Amiri Z, Shobeiri P, Malekpour MR, Saeedi Moghaddam S, Ghanbari A, et al. Burden of autism spectrum disorders in North Africa and Middle East from 1990 to 2019: A systematic analysis for the Global Burden of Disease Study 2019. Brain Behav. 2023;13(7):e3067.

2. Salari N, Rasoulpoor S, Rasoulpoor S, Shohaimi S, Jafarpour S, Abdoli N, et al. The global prevalence of autism spectrum disorder: a comprehensive systematic review and meta-analysis. Ital J Pediatr. 2022;48(1):112.

3. Hodges H, Fealko C, Soares N. Autism spectrum disorder: definition, epidemiology, causes, and clinical evaluation. Transl Pediatr. 2020;9(Suppl 1):S55–S65.

4. Yu Y, Ozonoff S, Miller M. Assessment of Autism Spectrum Disorder. Assessment. 2024;31(1):24–41.

5. World Health Organization. Autism 2023 [Available from: https://www.who.int/news-room/fact-sheets/detail/autism-spectrum-disorders.

6. Aylward BS, Gal-Szabo DE, Taraman S. Racial, Ethnic, and Sociodemographic Disparities in Diagnosis of Children with Autism Spectrum Disorder. J Dev Behav Pediatr. 2021;42(8):682–9.

7. Zeidan J, Fombonne E, Scorah J, Ibrahim A, Durkin MS, Saxena S, et al. Global prevalence of autism: A systematic review update. Autism Res. 2022;15(5):778–90.

8. Talantseva OI, Romanova RS, Shurdova EM, Dolgorukova TA, Sologub PS, Titova OS, et al. The global prevalence of autism spectrum disorder: A three-level meta-analysis. Front Psychiatry. 2023;14:1071181.

9. Bernardo WM. PRISMA statement and PROSPERO. Int Braz J Urol. 43. Brazil 2017. p. 383–4.

10. GA Wells BS, D O’Connell, J Peterson, V Welch, M Losos, P Tugwell. The Newcastle-Ottawa Scale (NOS) for assessing the quality of nonrandomised studies in meta-analyses. 2011.

11. Doi SA, Barendregt JJ, Khan S, Thalib L, Williams GM. Advances in the meta-analysis of heterogeneous clinical trials II: The quality effects model. Contemp Clin Trials. 2015;45(Pt A):123–9.

12. Higgins JP, Thompson SG, Deeks JJ, Altman DG. Measuring inconsistency in meta-analyses. BMJ. 2003;327(7414):557-60.

13. Furuya-Kanamori L, Barendregt JJ, Doi SAR. A new improved graphical and quantitative method for detecting bias in meta-analysis. Int J Evid Based Healthc. 2018;16(4):195–203.

14. Page MJ, Moher D, Bossuyt PM, Boutron I, Hoffmann TC, Mulrow CD, et al. PRISMA 2020 explanation and elaboration: updated guidance and exemplars for reporting systematic reviews. BMJ (Clinical research ed). 2021;372:n160.

15. Al-Farsi YM, Al-Sharbati MM, Al-Farsi OA, Al-Shafaee MS, Brooks DR, Waly MI. Brief report: Prevalence of autistic spectrum disorders in the Sultanate of Oman. J Autism Dev Disord. 2011;41(6):821–5.

16. Eapen V, Mabrouk AA, Zoubeidi T, Yunis F. Prevalence of pervasive developmental disorders in preschool children in the UAE. J Trop Pediatr. 2007;53(3):202–5.

17. Sasanfar R, Haddad SA, Tolouei A, Ghadami M, Yu D, Santangelo SL. Paternal age increases the risk for autism in an Iranian population sample. Mol Autism. 2010;1(1):2.

18. Al-Mamri W, Idris AB, Dakak S, Al-Shekaili M, Al-Harthi Z, Alnaamani AM, et al. Revisiting the Prevalence of Autism Spectrum Disorder among Omani Children: A multicentre study. Sultan Qaboos Univ Med J. 2019;19(4):e305–e9.

19. Alshaban F, Aldosari M, Al-Shammari H, El-Hag S, Ghazal I, Tolefat M, et al. Prevalence and correlates of autism spectrum disorder in Qatar: a national study. J Child Psychol Psychiatry. 2019;60(12):1254–68.

20. Chaaya M, Saab D, Maalouf FT, Boustany RM. Prevalence of Autism Spectrum Disorder in Nurseries in Lebanon: A Cross Sectional Study. J Autism Dev Disord. 2016;46(2):514–22.

21. Faraji Goodarzi M, Taee N, Abbasi Hormozi PJECD, Care. Evaluation of autistic spectrum disorders screening in children of Khorramabad (West of Iran) between 2015 and 2016. 2019;189:1509–14.

22. Manzouri L, Yousefian S, Keshtkari A, Hashemi N. Advanced Parental Age and Risk of Positive Autism Spectrum Disorders Screening. Int J Prev Med. 2019;10:135.

23. Mohammadi MR, Ahmadi N, Khaleghi A, Zarafshan H, Mostafavi SA, Kamali K, et al. Prevalence of Autism and its Comorbidities and the Relationship with Maternal Psychopathology: A National Population-Based Study. Arch Iran Med. 2019;22(10):546–53.

24. Sabbagh HJ, Al-Jabri BA, Alsulami MA, Hashem LA, Aljubour AA, Alamoudi RA. Prevalence and characteristics of autistic children attending autism centres in 2 major cities in Saudi Arabia: A cross-sectional study. Saudi Med J. 2021;42(4):419–27.

25. Samadi SA, Mahmoodizadeh A, McConkey R. A national study of the prevalence of autism among five-year-old children in Iran. Autism. 2012;16(1):5–14.

26. Samadi SA, McConkey R. Screening for Autism in Iranian Preschoolers: Contrasting M-CHAT and a Scale Developed in Iran. J Autism Dev Disord. 2015;45(9):2908–16.

27. Yousef AM, Roshdy EH, Abdel Fattah NR, Said RM, Atia MM, Hafez EM, et al. Prevalence and risk factors of autism spectrum disorders in preschool children in Sharkia, Egypt: a community-based study. Middle East Current Psychiatry. 2021;28(1):36.

28. Zeglam AM, Maound AJ. Prevalence of autistic spectrum disorders in Tripoli, Libya: the need for more research and planned services. East Mediterr Health J. 2012;18(2):184–8.

29. Al-Ansari AM, Ahmed MM. Epidemiology of autistic disorder in Bahrain: prevalence and obstetric and familial characteristics. East Mediterr Health J. 2013;19(9):769–74.

30. AlBatti TH, Alsaghan LB, Alsharif MF, Alharbi JS, BinOmair AI, Alghurair HA, et al. Prevalence of autism spectrum disorder among Saudi children between 2 and 4 years old in Riyadh. Asian J Psychiatr. 2022;71:103054.

31. Salhia HO, Al-Nasser LA, Taher LS, Al-Khathaami AM, El-Metwally AA. Systemic review of the epidemiology of autism in Arab Gulf countries. Neurosciences (Riyadh). 2014;19(4):291–6.

32. Samms-Vaughan ME. The status of early identification and early intervention in autism spectrum disorders in lower- and middle-income countries. Int J Speech Lang Pathol. 2014;16(1):30–5.

33. Rylaarsdam L, Guemez-Gamboa A. Genetic Causes and Modifiers of Autism Spectrum Disorder. Front Cell Neurosci. 2019;13:385.

34. Wang C, Geng H, Liu W, Zhang G. Prenatal, perinatal, and postnatal factors associated with autism: A meta-analysis. Medicine (Baltimore). 2017;96(18):e6696.

35. Mamidala MP, Kalikiri MK, Praveen Kumar PT, Rajesh N, Vallamkonda OR, Rajesh V. Consanguinity in India and its association with autism spectrum disorder. Autism Res. 2015;8(2):224–8.

36. Brynge M, Sjoqvist H, Gardner RM, Lee BK, Dalman C, Karlsson H. Maternal infection during pregnancy and likelihood of autism and intellectual disability in children in Sweden: a negative control and sibling comparison cohort study. Lancet Psychiatry. 2022;9(10):782–91.

37. Elsabbagh M, Divan G, Koh Y-J, Kim YS, Kauchali S, Marcín C, et al. Global Prevalence of Autism and Other Pervasive Developmental Disorders. 2012;5(3):160–79.

38. Doi SA, Thalib L. A quality-effects model for meta-analysis. Epidemiology. 2008;19(1):94–100.

