## Supplementary materials for "Estimates of the Prevalence of Autism Spectrum Disorder in the Middle East and North Africa Region: A Systematic Review and Meta-Analysis"

[2] Sidra Medicine, Doha, Qatar.

[3] Hamad Hospital, Doha, Qatar.

[4] Weill Cornell Medicine Qatar, Doha, Qatar.

[*] These authors contributed equally.

Corresponding and guarantor author:

Tawanda Chivese,

Department of Population Medicine, College of Medicine, QU Health, Qatar University, Doha, Qatar

P O BOX 2713

Doha, Qatar

Telephone number: +974 4403 7831

### PRISMA Checklist

Supplementary Table S1. PRISMA Checklist.

| Section and Topic | Item # | Checklist item | Location where item is reported (page) |
|---|---|---|---|
| **TITLE** | | | |
| Title | 1 | Identify the report as a systematic review. | 1 |
| **ABSTRACT** | | | |
| Abstract | 2 | See the PRISMA 2020 for Abstracts checklist. | 2 |
| **INTRODUCTION** | | | |
| Rationale | 3 | Describe the rationale for the review in the context of existing knowledge. | 3 – 4 |
| Objectives | 4 | Provide an explicit statement of the objective(s) or question(s) the review addresses. | 3 – 4 |
| **METHODS** | | | |
| Eligibility criteria | 5 | Specify the inclusion and exclusion criteria for the review and how studies were grouped for the syntheses. | 4 – 5 |
| Information sources | 6 | Specify all databases, registers, websites, organisations, reference lists and other sources searched or consulted to identify studies. Specify the date when each source was last searched or consulted. | 4 |
| Search strategy | 7 | Present the full search strategies for all databases, registers and websites, including any filters and limits used. | 4 |
| Selection process | 8 | Specify the methods used to decide whether a study met the inclusion criteria of the review, including how many reviewers screened each record and each report retrieved, whether they worked independently, and if applicable, details of automation tools used in the process. | 4 |
| Data collection process | 9 | Specify the methods used to collect data from reports, including how many reviewers collected data from each report, whether they worked independently, any processes for obtaining or confirming data from study investigators, and if applicable, details of automation tools used in the process. | 5 |
| Data items | 10a | List and define all outcomes for which data were sought. Specify whether all results that were compatible with each outcome domain in each study were sought (e.g. for all measures, time points, analyses), and if not, the methods used to decide which results to collect. | 5 |
| | 10b | List and define all other variables for which data were sought (e.g. participant and intervention characteristics, funding sources). Describe any assumptions made about any missing or unclear information. | 5 |
| Study risk of bias assessment | 11 | Specify the methods used to assess risk of bias in the included studies, including details of the tool(s) used, how many reviewers assessed each study and whether they worked independently, and if applicable, details of automation tools used in the process. | 5 |
| Effect measures | 12 | Specify for each outcome the effect measure(s) (e.g. risk ratio, mean difference) used in the synthesis or presentation of results. | 5 – 6 |
| Synthesis methods | 13a | Describe the processes used to decide which studies were eligible for each synthesis (e.g. tabulating the study intervention characteristics and comparing against the planned groups for each synthesis (item #5)). | 5 – 6 |
| | 13b | Describe any methods required to prepare the data for presentation or synthesis, such as handling of missing summary statistics, or data conversions. | 5 – 6 |
| | 13c | Describe any methods used to tabulate or visually display results of individual studies and syntheses. | 5 – 6 |
| | 13d | Describe any methods used to synthesize results and provide a rationale for the choice(s). If meta-analysis was performed, describe the model(s), method(s) to identify the presence and extent of statistical heterogeneity, and software package(s) used. | 5 – 6 |

| Section and Topic | Item # | Checklist item | Location where item is reported (page) |
|---|---|---|---|
| | 13e | Describe any methods used to explore possible causes of heterogeneity among study results (e.g. subgroup analysis, meta-regression). | 6 |
| | 13f | Describe any sensitivity analyses conducted to assess robustness of the synthesized results. | 6 |
| Reporting bias assessment | 14 | Describe any methods used to assess risk of bias due to missing results in a synthesis (arising from reporting biases). | 5 – 6 |
| Certainty assessment | 15 | Describe any methods used to assess certainty (or confidence) in the body of evidence for an outcome. | N/A |
| **RESULTS** | | | |
| Study selection | 16a | Describe the results of the search and selection process, from the number of records identified in the search to the number of studies included in the review, ideally using a flow diagram. | 6 |
| | 16b | Cite studies that might appear to meet the inclusion criteria, but which were excluded, and explain why they were excluded. | 6 – 7 |
| Study characteristics | 17 | Cite each included study and present its characteristics. | 7 – 10 |
| Risk of bias in studies | 18 | Present assessments of risk of bias for each included study. | 11 |
| Results of individual studies | 19 | For all outcomes, present, for each study: (a) summary statistics for each group (where appropriate) and (b) an effect estimate and its precision (e.g. confidence/credible interval), ideally using structured tables or plots. | 11 – 13 |
| Results of syntheses | 20a | For each synthesis, briefly summarise the characteristics and risk of bias among contributing studies. | 11 – 13 |
| | 20b | Present results of all statistical syntheses conducted. If meta-analysis was done, present for each the summary estimate and its precision (e.g. confidence/credible interval) and measures of statistical heterogeneity. If comparing groups, describe the direction of the effect. | 11 – 13 |
| | 20c | Present results of all investigations of possible causes of heterogeneity among study results. | 11 – 13 |
| | 20d | Present results of all sensitivity analyses conducted to assess the robustness of the synthesized results. | 11 – 13 |
| Reporting biases | 21 | Present assessments of risk of bias due to missing results (arising from reporting biases) for each synthesis assessed. | 11 – 13 |
| Certainty of evidence | 22 | Present assessments of certainty (or confidence) in the body of evidence for each outcome assessed. | N/A |
| **DISCUSSION** | | | |
| Discussion | 23a | Provide a general interpretation of the results in the context of other evidence. | 14 -15 |
| | 23b | Discuss any limitations of the evidence included in the review. | 14-15 |
| | 23c | Discuss any limitations of the review processes used. | 15 |
| | 23d | Discuss implications of the results for practice, policy, and future research. | 15 |
| **OTHER INFORMATION** | | | |
| Registration and protocol | 24a | Provide registration information for the review, including register name and registration number, or state that the review was not registered. | 4 |
| | 24b | Indicate where the review protocol can be accessed, or state that a protocol was not prepared. | 4 |
| | 24c | Describe and explain any amendments to information provided at registration or in the protocol. | N/A |

| Section and Topic | Item # | Checklist item | Location where item is reported (page) |
|---|---|---|---|
| Support | 25 | Describe sources of financial or non-financial support for the review, and the role of the funders or sponsors in the review. | 17 |
| Competing interests | 26 | Declare any competing interests of review authors. | 17 |
| Availability of data, code and other materials | 27 | Report which of the following are publicly available and where they can be found: template data collection forms; data extracted from included studies; data used for all analyses; analytic code; any other materials used in the review. | 18 |

**Search Strategy**

Supplementary Table S2. PubMed Medline Search. Results: 703. Last updated: 20 January 2024.

| |
|---|
| ("Autism Spectrum Disorder"[Mesh] OR "Autistic Disorder"[Mesh] OR "Asperger Syndrome"[Mesh] OR "autism spectrum disorder" [tiab] OR "ASD" [title] OR "autism spectrum condition" [tiab] OR "ASC" [title] OR "Autism" [tiab] OR "Kanner syndrome" [tiab] OR "autistic disorder" [tiab] OR "childhood autism" [tiab] OR "Asperger syndrome" [tiab] OR "childhood disintegrative disorder" [tiab] OR "pervasive developmental disorder not otherwise specified" [tiab])<br>AND<br>("MENA" [tiab] OR "Middle East*" [tiab] OR "North Africa*" [tiab] OR "Algeria*" [tiab] OR Bahrain* [tiab] OR Djibouti* [tiab] OR Egypt* [tiab] OR Iran* [tiab] OR Iraq* [tiab] OR Jordan* [tiab] OR Kuwait* [tiab] OR Leban* [tiab] OR Liby* [tiab] OR Malt* [tiab] OR Mauritania* [tiab] OR Morocc* [tiab] OR Oman* [tiab] OR Palestin* [tiab] OR Qatar* [tiab] OR "Saudi" [tiab] OR Somalia* [tiab] OR Sudan* [tiab] OR Syria* [tiab] OR Tunisia* [tiab] OR "United Arab Emirates" [tiab] OR UAE [title] OR Emirati [tiab] OR Yemen* [tiab]) |

Supplementary Table S3. Embase Search. Results: 949. Last updated: 20 January 2024.

| |
|---|
| ('Autism Spectrum Disorder':ti,ab OR 'Autistic Disorder':ti,ab OR 'Asperger Syndrome':ti,ab OR ASD:ti OR 'autism spectrum condition':ti,ab OR ASC:ti OR Autism:ti,ab OR 'Kanner syndrome':ti,ab OR 'childhood autism':ti,ab OR 'childhood disintegrative disorder':ti,ab OR 'pervasive developmental disorder not otherwise specified':ti,ab)<br>AND<br>(MENA:ti,ab OR 'Middle East*':ti,ab OR 'North Africa*':ti,ab OR Algeria*:ti,ab OR Bahrain*:ti,ab OR Djibouti*:ti,ab OR Egypt*:ti,ab OR Iran*:ti,ab OR Iraq*:ti,ab OR Jordan*:ti,ab OR Kuwait*:ti,ab OR Leban*:ti,ab OR Liby*:ti,ab OR Malt*:ti,ab OR Mauritania*:ti,ab OR Morocc*:ti,ab OR Oman*:ti,ab OR Palestin*:ti,ab OR Qatar*:ti,ab OR Saudi:ti,ab OR Somalia*:ti,ab OR Sudan*:ti,ab OR Syria*:ti,ab OR Tunisia*:ti,ab OR 'United Arab Emirates':ti,ab OR UAE:ti OR Emirati:ti,ab OR Yemen*:ti,ab) |

Supplementary Table S4. Scopus Search. Results: 1118. Last updated: 20 January 2024.

| |
|---|
| (TITLE-ABS("autism spectrum disorder") OR TITLE(ASD) OR TITLE-ABS("autism spectrum condition") OR TITLE(ASC) OR TITLE-ABS(Autism) OR TITLE-ABS("Kanner syndrome") OR TITLE-ABS("autistic disorder") OR TITLE-ABS("childhood autism") OR TITLE-ABS("Asperger syndrome") OR TITLE-ABS("childhood disintegrative disorder") OR TITLE-ABS("pervasive developmental disorder not otherwise specified"))<br>AND<br>(TITLE-ABS(MENA) OR TITLE-ABS("Middle East*") OR TITLE-ABS("North Africa*") OR TITLE-ABS(Algeria*) OR TITLE-ABS(Bahrain*) OR TITLE-ABS(Djibouti*) OR TITLE-ABS(Egypt*) OR TITLE-ABS(Iran*) OR TITLE-ABS(Iraq*) OR TITLE-ABS(Jordan*) OR TITLE-ABS(Kuwait*) OR TITLE-ABS(Leban*) OR TITLE-ABS(Liby*) OR TITLE-ABS(Malt*) OR TITLE-ABS(Mauritania*) OR TITLE-ABS(Morocc*) OR TITLE-ABS(Oman*) OR TITLE-ABS(Palestin*) OR TITLE-ABS(Qatar*) OR TITLE-ABS(Saudi) OR TITLE-ABS(Somalia*) OR TITLE-ABS(Sudan*) OR TITLE-ABS(Syria*) OR TITLE-ABS(Tunisia*) OR TITLE-ABS("United Arab Emirates") OR TITLE(UAE) OR TITLE-ABS(Emirati) OR TITLE-ABS(Yemen*)) |

Supplementary Table S5. CINHAL. Results: 302. Last updated: 20 January 2024.

| |
|---|
| ((TI "Autism Spectrum Disorder" OR AB "Autism Spectrum Disorder") OR (TI "Autistic Disorder" OR AB "Autistic Disorder") OR (TI "Asperger Syndrome" OR AB "Asperger Syndrome") OR (TI ASD) OR (TI "autism spectrum condition" OR AB "autism spectrum condition") OR (TI ASC) OR (TI Autism OR AB Autism) OR (TI "Kanner syndrome" OR AB "Kanner syndrome") OR (TI "childhood autism" OR AB "childhood autism") OR (TI "childhood disintegrative disorder" OR AB "childhood disintegrative disorder") OR (TI "pervasive developmental disorder not otherwise specified" OR AB "pervasive developmental disorder not otherwise specified"))<br>AND<br>((TI MENA OR AB MENA) OR (TI "Middle East*" OR AB "Middle East*") OR (TI "North Africa*" OR AB "North Africa*") OR (TI Algeria* OR AB Algeria*) OR (TI Bahrain* OR AB Bahrain*) OR (TI Djibouti* OR AB Djibouti*) OR (TI Egypt* OR AB Egypt*) OR (TI Iran* OR AB Iran*) OR (TI Iraq* OR AB Iraq*) OR (TI Jordan* OR AB Jordan*) OR (TI |

Kuwait* OR AB Kuwait*) OR (TI Leban* OR AB Leban*) OR (TI Liby* OR AB Liby*) OR (TI Malt* OR AB Malt*) OR (TI Mauritania* OR AB Mauritania*) OR (TI Morocc* OR AB Morocc*) OR (TI Oman* OR AB Oman*) OR (TI Palestin* OR AB Palestin*) OR (TI Qatar* OR AB Qatar*) OR (TI Saudi OR AB Saudi) OR (TI Somalia* OR AB Somalia*) OR (TI Sudan* OR AB Sudan*) OR (TI Syria* OR AB Syria*) OR (TI Tunisia* OR AB Tunisia*) OR (TI "United Arab Emirates" OR AB "United Arab Emirates") OR (TI UAE) OR (TI Emirati OR AB Emirati) OR (TI Yemen* OR AB Yemen*))

### Quality Assessment Tools

Supplementary Table S6. Newcastle-Ottawa quality assessment scale for case control studies.

**Selection**

A. Is the case defintion adequate?
   2. yes, with independent validation
   1. yes, eg record linkage or based on self reports
   0. no description

B. Representitiveness of the cases
   1. consecutive or obviously representative series of cases
   0. potential for selection biases or not stated

C. Selection of controls
   2. community controls
   1. hospital controls
   0. no description

D. Definition of controls
   1. no history of disease (endpoint)
   0. no description of source

**Comparability**

E. Comparability of cases and controls for age and sex
   1. Yes
   0. No description

F. Comparability of cases and controls for other variables
   1. Yes
   0. No description

**Exposure**

G. Ascertainment of exposure
   4. secure record (eg surgical records)
   3. structured interview where blind to case/control status
   2. interview not blinded to case/control status
   1. written self report or medical record only
   0. no description

H. Same method of ascertainment for cases and controls
   1. Yes
   0. No

I. Non-response rate
   2. same rate for both groups
   1. non respondents described
   0. rate different and no designation

Supplementary Table S7. Adapted Newcastle-Ottawa quality assessment scale for cross sectional studies.

**Selection**

A. Representitiveness of the sample
   3. truly representitive of the average population (all subjects or random sample)
   2. somewhat representitive of the average population (nonrandom sample)
   1. selected group of users, eg. nurses, volunteers
   0. no description

B. Sample size
   1. Justified and satisfactory (including sample size calculation).
   0. Not justified
   0. no description of sample size

C. Non-respondants
   2. Comparability between respondants and non-respondants characteristics is established, and the respons rate is satisfactory
   1. Response rate is unsatisfactory, or the comparability is unsatisfactory
   0. no description of the response rate or comparability of characteristics

D. Ascertainment of exposure (risk factor)
   2. Validated measurement tool
   1. Non-validated measurement tool, but the tool is available or described
   0. no description of the measurement tool

**Comparability**

E. Comparability of both groups for age and sex
   1. Yes
   0. No description

F. Comparability of both groups for other variables
   1. Yes
   0. No description

**Outcome**

G. Assessment of outcome
   3. independent blind assessment
   2. record linkage
   1. self report
   0. no description

H. Statistical test
   1. The statistical test used to analyze the data is clearly described and appropriate, and the measurement of the association is presented, including confidence intervals and the probability level (p value)
   0. The statistical test is not appropriate, not described or incomplete

**Excluded Reports**

Supplementary Table S8. Excluded reports at full-text screening stage with the reasons for exclusion.

| First Author | Year | Title | Reason for Exclusion |
|---|---|---|---|
| Eapen (1) | 2003 | Epidemiological study of developmental and behavioural disorders in three year old children: A UAE study | Full-text not retrieved |
| Al-Ayadhi (2) | 2005 | Heavy metals and trace elements in hair samples of autistic children in central Saudi Arabia | Wrong outcome |
| Al-Ayadhi (3) | 2005 | Pro-inflammatory cytokines in autistic children in central Saudi Arabia | Wrong outcome |
| Al-Ayadhi (4) | 2005 | Autoimmune connection of autism in Central Saudi Arabia | Wrong outcome |
| Al-Ayadhi (5) | 2005 | Altered oxytocin and vasopressin levels in autistic children in Central Saudi Arabia | Wrong outcome |
| Mostafa (6) | 2008 | Serum anti-myelin - Associated glycoprotein antibodies in Egyptian Autistic children | Wrong outcome |
| Al-Salehi (7) | 2009 | Autism in Saudi Arabia: PresentatioN, Clinical Correlates and Comorbidity | Wrong publication type |
| Walsh (8) | 2010 | High-throughput dna sequencing in autism spectrum disorders (ASD) | Wrong publication type |
| Meguid (9) | 2010 | Reduced serum levels of 25-hydroxy and 1,25-dihydroxy vitamin D in Egyptian children with autism | Wrong outcome |
| El-Ansary (10) | 2010 | Measurement of selected ions related to oxidative stress and energy metabolism in Saudi autistic children | Wrong outcome |
| Al-Farsi (11) | 2011 | Malnutrition among preschool-aged autistic children in Oman | Wrong outcome |
| Al-Yafee (12) | 2011 | Novel metabolic biomarkers related to sulfur-dependent detoxification pathways in autistic patients of Saudi Arabia | Wrong outcome |
| El-Ansary (13) | 2011 | Proinflammatory and proapoptotic markers in relation to mono and di-cations in plasma of autistic patients from Saudi Arabia | Wrong outcome |
| Hussein (14) | 2011 | Characteristics of autism spectrum disorders in a sample of egyptian and saudi patients: Transcultural cross sectional study | Wrong outcome |
| Meguid (15) | 2011 | Evaluation of oxidative stress in autism: Defective antioxidant enzymes and increased lipid peroxidation | Wrong outcome |
| Mohareri (16) | 2011 | Attention deficit hyperactivity symptoms in children with autistic spectrum disorder | Wrong outcome |

| | | | |
|---|---|---|---|
| Abd Elhameed (17) | 2011 | A controlled study of the risk factors and clinical picture of children with Autism in an Egyptian sample | Full-text not retrieved |
| El-Baz (18) | 2011 | Risk factors for autism: An Egyptian study | Wrong outcome |
| Waly (19) | 2011 | Low serum levels of glutathione, homocysteine and total antioxidant capacity are associated with increased risk of autism in Oman | Wrong outcome |
| Amr (20) | 2011 | Sex differences in Arab children with Autism spectrum disorders | Wrong outcome |
| Mohammadi (21) | 2011 | Autism spectrum disorders in Iran | Wrong publication type |
| Masri (22) | 2011 | Profile of developmental delay in children under five years of age in a highly consanguineous community: A hospital-based study - Jordan | Wrong outcome |
| Gebril (23) | 2011 | HFE gene polymorphisms and the risk for autism in Egyptian children and impact on the effect of oxidative stress | Wrong outcome |
| El-Ansary (24) | 2011 | Impaired plasma phospholipids and relative amounts of essential polyunsaturated fatty acids in autistic patients from Saudi Arabia | Wrong outcome |
| Zeglam (25) | 2012 | Is there a need for a focused health care service for children with autistic spectrum disorders? A keyhole look at this problem in Tripoli, Libya | Wrong publication type |
| El-Ansary (26) | 2012 | Lipid mediators in plasma of autism spectrum disorders | Wrong outcome |
| El-Ansary (27) | 2012 | Relationship between chronic lead toxicity and plasma neurotransmitters in autistic patients from Saudi Arabia | Wrong outcome |
| Essa (28) | 2012 | Increased markers of oxidative stress in autistic children of the Sultanate of Oman | Wrong outcome |
| Elshahawi (29) | 2012 | Possible association of certain HLA-DRB1 alleles with autism in Egyptian children: Relation to family history of autoimmunity | Wrong publication type |
| Zakareia (30) | 2012 | Study of dual angiogenic/neurogenic growth factors among Saudi autistic children and their correlation with the severity of this disorder | Wrong outcome |
| Amr (31) | 2012 | Comorbid psychiatric disorders in Arab children with Autism spectrum disorders | Wrong outcome |
| Al-Sharbati (32) | 2012 | Autistic Spectrum Disorder (ASD) Among Omani Children Below 6 Years: A Five Year Retrospective Descriptive Study | Wrong publication type |
| Al-Farsi (33) | 2013 | Impact of nutrition on serum levels of docosahexaenoic acid among Omani children with autism | Wrong outcome |
| Al-Farsi (34) | 2013 | Levels of heavy metals and essential minerals in hair samples of children with autism in Oman: A case-control study | Wrong outcome |

| | | | |
|---|---|---|---|
| Al-Rubaye (35) | 2013 | Purine metabolism and oxidative stress in children with autistic spectrum disorders | Wrong outcome |
| Zakareia (36) | 2013 | Evaluation of plasma soluble fatty acid synthase levels among Saudi autistic children: Relation to disease severity | Full-text not retrieved |
| Al-Farsi (37) | 2013 | Association of gestational diabetes mellitus with occurrence of Autism: A cohort study | Wrong publication type |
| Bashir (38) | 2013 | Serum level of desert hedgehog protein in autism spectrum disorder: Preliminary results | Wrong outcome |
| Hamadé (39) | 2013 | Autism in children and correlates in lebanon: A pilot case-control study | Wrong outcome |
| Hashim (40) | 2013 | Association between plasma levels of transforming growth factor-β1, IL-23 and IL-17 and the severity of autism in Egyptian children | Wrong outcome |
| Mousavizadeh (41) | 2013 | Association of human mtDNA mutations with autism in Iranian patients | Wrong publication type |
| Kaddah (42) | 2013 | Screening for autism in low-birth-weight Egyptian toddlers | Wrong population |
| Al-Hakbany (43) | 2014 | The Relationship of HLA Class I and II Alleles and Haplotypes with Autism: A Case Control Study | Wrong outcome |
| Ranjbar (44) | 2014 | Comparison of urinary oxidative biomarkers in Iranian children with autism | Wrong outcome |
| Hodgson (45) | 2014 | Decreased glutathione and elevated hair mercury levels are associated with nutritional deficiency-based autism in Oman | Wrong outcome |
| Yassa (46) | 2014 | Autism: A form of lead and mercury toxicity | Wrong outcome |
| Alabdali (47) | 2014 | Association of social and cognitive impairment and biomarkers in autism spectrum disorders | Wrong outcome |
| El-Ansary (48) | 2014 | Role of amino acids in the pathophysiology of autism spectrum disorders in Saudi and Egyptian population samples | Wrong outcome |
| Shawky (49) | 2014 | Study of genotype-phenotype correlation of methylene tetrahydrofolate reductase (MTHFR) gene polymorphisms in a sample of Egyptian autistic children | Wrong outcome |
| Afsharpaiman (50) | 2014 | An assessment of toxoplasmosis antibodies seropositivity in children suffering autism | Wrong outcome |
| Mohamed (51) | 2015 | Assessment of Hair Aluminum, Lead, and Mercury in a Sample of Autistic Egyptian Children: Environmental Risk Factors of Heavy Metals in Autism | Wrong outcome |
| Halepoto (52) | 2015 | Correlation Between Hedgehog (Hh) Protein Family and Brain-Derived Neurotrophic Factor (BDNF) in Autism Spectrum Disorder (ASD) | Wrong outcome |

| | | | |
|---|---|---|---|
| Fanid (53) | 2015 | Association between common single- nucleotide polymorphism of reelin gene, rs736707 (C/T) with autism spectrum disorder in Iranian-Azeri patients | Wrong outcome |
| Fernell (54) | 2015 | Autism spectrum disorder and low vitamin D at birth: A sibling control study | Wrong population |
| Slama (55) | 2015 | Family history of psychiatric disorder and autism spectrum disorders: A study about 790 cases | Wrong publication type |
| Meguid (56) | 2015 | Evaluation of MTHFR genetic polymorphism as a risk factor in Egyptian autistic children and mothers | Wrong outcome |
| Mousavi (57) | 2015 | RoRa gene contribution to autism; another epigenetic layer on autism complexity | Wrong outcome |
| Ouhtit (58) | 2015 | Underlying factors behind the low prevalence of autism spectrum disorders in oman sociocultural perspective | Wrong publication type |
| Yavarna (59) | 2015 | High diagnostic yield of clinical exome sequencing in Middle Eastern patients with Mendelian disorders | Wrong outcome |
| Khakzad (60) | 2015 | Transforming growth factor beta 1 869T/C and 915G/C polymorphisms and risk of autism spectrum disorders | Wrong outcome |
| Afrazeh (61) | 2015 | Measurement of Serum Superoxide Dismutase and Its Relevance to Disease Intensity Autistic Children | Wrong outcome |
| Saad (62) | 2015 | ADHD, autism and neuroradiological complications among phenylketonuric children in Upper Egypt | Wrong outcome |
| Al-Sharbati (63) | 2016 | Profile of mental and behavioral disorders among preschoolers in a tertiary care hospital in Oman: A retrospective study | Wrong outcome |
| Fahmy (64) | 2016 | Vitamin D intake and sun exposure in autistic children | Wrong outcome |
| Haghiri (65) | 2016 | Analysis of methionine synthase (rs1805087) gene polymorphism in autism patients in northern Iran | Wrong outcome |
| El-Ansary (66) | 2016 | Data of multiple regressions analysis between selected biomarkers related to glutamate excitotoxicity and oxidative stress in Saudi autistic patients | Wrong outcome |
| No author name (67) | 2016 | Environmental toxic pollutant and trace elements in Egyptian children with autism | Full-text not retrieved |
| Dinkler (68) | 2016 | Maltreatment-associated neurodevelopmental problems: Environment and genetics | Wrong population |
| Elhawary (69) | 2016 | Vulnerability of genetic variants to the risk of autism among Saudi children | Wrong outcome |
| Fanid (70) | 2016 | An Association Analysis of Reelin Gene (RELN) exon 22 (G/C), Rs.362691, polymorphism with | Wrong outcome |

| | | | |
|---|---|---|---|
| | | autism spectrum disorder among Iranian-Azeri population | |
| Khaled (71) | 2016 | Altered urinary porphyrins and mercury exposure as biomarkers for autism severity in Egyptian children with autism spectrum disorder | Wrong outcome |
| Meguid (72) | 2016 | Impact of oxidative stress on autism spectrum disorder behaviors in children with autism | Full-text not retrieved |
| Mohammed (73) | 2016 | Incidence of autism in high risk neonatal follow up | Wrong population |
| Alhowikan (74) | 2017 | Secretagogin (SCGN) plasma levels and their association with cognitive and social behavior in children with autism spectrum disorder (ASD) | Wrong outcome |
| Alshaban (75) | 2017 | Autism spectrum disorder in Qatar: Profiles and correlates of a large clinical sample | Wrong outcome |
| Bener (76) | 2017 | Iron and Vitamin D levels among autism spectrum disorders children | Wrong outcome |
| Desoky (77) | 2017 | Biochemical assessments of thyroid profile, serum 25-hydroxycholecalciferol and cluster of differentiation 5 expression levels among children with autism | Wrong outcome |
| Ashaat (78) | 2017 | Altered adaptive cellular immune function in a group of Egyptian children with autism | Wrong outcome |
| Firouzabadi (79) | 2017 | Copy Number Variants in Patients with Autism and Additional Clinical Features: Report of VIPR2 Duplication and a Novel Microduplication Syndrome | Wrong outcome |
| Khaniani (80) | 2017 | Autistic Phenotype of Permutation and Intermediate Alleles of FMR1 Gene | Wrong outcome |
| Kourtian (81) | 2017 | Candidate Genes for Inherited Autism Susceptibility in the Lebanese Population | Wrong outcome |
| Zeglam (82) | 2017 | Epidemiology of autism in Libya: Uncovering of the first autism findings | Wrong publication type |
| Dinkler (83) | 2017 | Maltreatment-associated neurodevelopmental disorders: a co-twin control analysis | Wrong population |
| Ajabi (84) | 2017 | A study of MTRR 66A>G gene polymorphism in patients with autism from northern Iran | Wrong outcome |
| Hosseinpour (85) | 2017 | Neuropilin-2 rs849563 gene variations and susceptibility to autism in Iranian population: A case-control study | Wrong outcome |
| Hamedani (86) | 2017 | Ras-like without CAAX 2 (RIT2): a susceptibility gene for autism spectrum disorder | Wrong outcome |
| El-Ansary (87) | 2017 | Relationship between selenium, lead, and mercury in red blood cells of Saudi autistic children | Wrong outcome |
| Noroozi (88) | 2017 | Association study of the vesicular monoamine transporter 1 (VMAT1) gene with autism in an Iranian population | Wrong outcome |

| | | | |
|---|---|---|---|
| Safari (89) | 2017 | Synaptosome-Associated Protein 25 (SNAP25) Gene Association Analysis Revealed Risk Variants for ASD, in Iranian Population | Wrong outcome |
| Sayad (90) | 2017 | Retinoic acid-related orphan receptor alpha (RORA) variants are associated with autism spectrum disorder | Wrong outcome |
| Zare (91) | 2017 | The association of CNTNAP2 rs7794745 gene polymorphism and autism in Iranian population | Wrong outcome |
| Zeglam (92) | 2017 | Early TV viewing and autistic spectrum disorder; A plausible hypothesis that should not be dismissed "Libyan Viewpoint" | Wrong publication type |
| Abdulamir (93) | 2018 | Acetylserotonin O-Methyltransferase (ASMT)/rs4446909 Polymorphism in Iraqi Autistic Children | Wrong outcome |
| Kadhim (94) | 2018 | Sero-positivity rate of rubella antibodies in Iraqi autistic children | Wrong outcome |
| Belkady (95) | 2018 | Chromosomal Abnormalities in Patients with Intellectual Disability: A 21-Year Retrospective Study | Wrong outcome |
| Delgado (96) | 2018 | Role of Metal Ion Dyshomeostasis in ASD: Evaluation of Copper, Zinc, and Selenium Levels in the North American ASD Population | Wrong publication type |
| Alshiban (97) | 2018 | Risk factors for Autism Spectrum Disorder (ASD) in Saudi Arabia | Wrong publication type |
| Mousavi (98) | 2018 | Autism and probable prerequisites: Severe and scheduled prenatal stresses at spotlight | Wrong outcome |
| Yousefian (99) | 2018 | Long-term exposure to ambient air pollution and autism spectrum disorder in children: A case-control study in Tehran, Iran | Wrong outcome |
| John (100) | 2018 | Is the prevalence of autism spectrum disorder decreased in black and ethnic children and adolescents? | Wrong publication type |
| Abdulamir (101) | 2018 | Serotonin and serotonin transporter levels in autistic children | Wrong outcome |
| Meguid (102) | 2018 | Frequency of risk factors and coexisting abnormalities in a population of Egyptian children with autism spectrum disorder | Wrong outcome |
| Olusanya (103) | 2018 | Developmental disabilities among children younger than 5 years in 195 countries and territories, 1990–2016: a systematic analysis for the Global Burden of Disease Study 2016 | Wrong population |
| Altamimi (104) | 2018 | Could Autism Be Associated With Nutritional Status in the Palestinian population? The Outcomes of the Palestinian Micronutrient Survey | Wrong study design |
| Noroozi (105) | 2018 | Association analysis of the GABRB3 promoter variant and susceptibility to autism spectrum disorder | Wrong outcome |

| | | | |
|---|---|---|---|
| Qasem (106) | 2018 | Impaired lipid metabolism markers to assess the risk of neuroinflammation in autism spectrum disorder | Wrong outcome |
| Arastoo (107) | 2018 | Evaluation of serum 25-Hydroxy vitamin D levels in children with autism spectrum disorder | Wrong outcome |
| Jabbar (108) | 2018 | Study of polymorphism in methionine synthase gene by RFLP-PCR in middle euphrates region of Iraq | Wrong outcome |
| Eftekharian (109) | 2018 | Expression Analysis of Protein Inhibitor of Activated STAT (PIAS) Genes in Autistic Patients | Wrong outcome |
| Sayad (110) | 2018 | Association of HLA alleles with autism | Wrong outcome |
| Guisso (111) | 2018 | Association of Autism with Maternal Infections, Perinatal and Other Risk Factors: A Case-Control Study | Wrong outcome |
| Oommen (112) | 2018 | Role of environmental factors in autism spectrum disorders in Saudi children aged 3-10 years in the Northern and Eastern regions of Saudi Arabia | Wrong outcome |
| Alawad (113) | 2019 | Lead among children with autism in Iraq. Is it a potential factor?: Lead level in Iraqi children with autism | Wrong outcome |
| Alzghoul (114) | 2019 | The association between levels of inflammatory markers in autistic children compared to their unaffected siblings and unrelated healthy controls | Wrong outcome |
| Elsayed (115) | 2019 | Study of autistic features among children and adolescents with congenital adrenal hyperplasia | Wrong population |
| Jabbar (116) | 2019 | Evaluation the Relationship Between DRD1 RS4532 Gene Polymorphisms and Autism by RFLP-PCR in Middle Euphrates Region of Iraq | Wrong outcome |
| Alhader (117) | 2019 | The potential interactive role of leptin with other hormones in the pathophysiology of asd in jordanian male children | Wrong publication type |
| Arab (118) | 2019 | Methylenetetrahydrofolate reductase gene variants confer potential vulnerability to autism spectrum disorder in a Saudi community | Wrong outcome |
| El Khatib (119) | 2019 | Gastrointestinal symptoms in children with autism spectrum disorders and correlation with autism functionality and severity: a case-control study | Wrong publication type |
| Ismail (120) | 2019 | Study of C677T variant of methylene tetrahydrofolate reductase gene in autistic spectrum disorder Egyptian children | Wrong outcome |
| Sayad (121) | 2019 | Association study of sequence variants in voltage-gated Ca2+ channel subunit alpha-1C and autism spectrum disorders | Wrong outcome |
| Bordeleau (122) | 2019 | Microglia as a potential link between pathological myelination and stereotypic behavior after exposure to maternal high-fat diet | Wrong publication type |

| | | | |
|---|---|---|---|
| Gleeson (123) | 2019 | The genetic and molecular basis of neurodevelopmental disorders | Wrong publication type |
| Hasan (124) | 2019 | Assessment of children upon suffering from withdrawals signs in Baghdad city, Iraq | Wrong outcome |
| Malek (125) | 2019 | Risk factors for autistic disorder: A case-control study | Wrong outcome |
| Sadek (126) | 2019 | Factors associated with autism in Saudi Arabia: A case-control study | Wrong outcome |
| Al-Zalabani (127) | 2019 | Is cesarean section delivery associated with autism spectrum disorder? | Wrong outcome |
| Alzghoul (128) | 2020 | The Association Between Serum Vitamin D3 Levels and Autism Among Jordanian Boys | Wrong outcome |
| Shamsedine (129) | 2020 | Breastfeeding association with autism spectrum disorders: A case-control study from Lebanon | Wrong outcome |
| Hussien (130) | 2020 | Evaluation of lead, copper and zinc levels for autistic children in thi-qar Province/Iraq | Wrong outcome |
| Khalil (131) | 2020 | Assessing risk factors of Autism Spectrum Disorders (ASD) and Attention Deficit Hyperactivity Disorder (ADHD) among Saudi Mothers: A retrospective study | Wrong publication type |
| Mossa (132) | 2020 | Evaluation of serum oxytocin hormone level in children with autism | Wrong outcome |
| Jenabi (133) | 2020 | Association between assisted reproductive technology and autism spectrum disorders in iran: A case-control study | Wrong outcome |
| Razjouyan (134) | 2020 | A Study of the Prevalence of Risk Factors Associated with Autism Spectrum Disorder among Affected Individuals in Tehran | Wrong comparison |
| Richa (135) | 2020 | Estimating the prevalence of autism spectrum disorder in Lebanon | Wrong outcome |
| Virolainen (136) | 2020 | Autism spectrum disorder in the United Arab Emirates: Potential environmental links | Wrong publication type |
| Muftin (137) | 2020 | Identification of MTHFR genetic polymorphism in Iraqi autistic children | Wrong outcome |
| Beiranvandi (138) | 2020 | The association of CNTNAP2 rs2710102 and ENGRAILED-2 rs1861972 genes polymorphism and autism in Iranian population | Wrong outcome |
| Darvish (139) | 2020 | Association of rs3735025 and rs9656169 variants with autism, and schizophrenia: A GWAS-replication study in an Iranian population | Wrong outcome |
| Kandeel (140) | 2020 | Impact of Clostridium Bacteria in Children with Autism Spectrum Disorder and Their Anthropometric Measurements | Wrong outcome |
| Mobasheri (141) | 2020 | Association between vitamin D receptor gene FokI and TaqI variants with autism spectrum disorder predisposition in Iranian population | Wrong outcome |

| | | | |
|---|---|---|---|
| Saad (142) | 2020 | Polymorphism of interleukin-1β and interleukin-1 receptor antagonist genes in children with autism spectrum disorders | Wrong outcome |
| Safari (143) | 2020 | The rs12826786 in HOTAIR lncRNA Is Associated with Risk of Autism Spectrum Disorder | Wrong outcome |
| Taheri (144) | 2020 | The rs594445 in MOCOS gene is associated with risk of autism spectrum disorder | Wrong outcome |
| Al-Bazzaz (145) | 2020 | Estimation of fasting serum levels of glucose, zinc, copper, zinc /copper ratio and their relation to the measured lipid profile in autistic patients and non-autistic controls in Jordan | Wrong outcome |
| Chehbani (146) | 2020 | The status of chemical elements in the blood plasma of children with autism spectrum disorder in Tunisia: a case-control study | Wrong outcome |
| Gerges (147) | 2020 | Risk and protective factors in autism spectrum disorders: A case control study in the lebanese population | Wrong outcome |
| Soliman (148) | 2020 | Autistic traits among Saudi university students; relations with emotional intelligence and alexithymia | Wrong publication type |
| Al-Mamari (149) | 2021 | Parental age and the risk of autism spectrum disorder in Oman a case-control study | Wrong outcome |
| Alomar (150) | 2021 | Vitamin D deficient diet and autism | Wrong outcome |
| Al-Ali (151) | 2021 | Determination of environmental risk factors of Autism in Kerbala city / Iraq 2020 | Wrong outcome |
| Al-Sarraj (152) | 2021 | Family-based genome-wide association study of autism spectrum disorder in middle eastern families | Wrong outcome |
| Hegazy (153) | 2021 | Environmental risk factors associated with children autism spectrum disorders in, menoufia governorate | Wrong outcome |
| Mondal (154) | 2021 | Role of glucose 6-phosphate dehydrogenase (G6PD) deficiency and its association to Autism Spectrum Disorders | Wrong outcome |
| Nawaz (155) | 2021 | Low Birth Weight Prevalence in Children Diagnosed with Neurodevelopmental Disorders in Dubai | Wrong outcome |
| Al Malki (156) | 2021 | Maternal toxoplasmosis and the risk of childhood autism: serological and molecular small-scale studies | Wrong outcome |
| Meguid (157) | 2021 | Awareness and risk factors of autism spectrum disorder in an Egyptian population | Wrong comparison |
| Rahmani (158) | 2021 | Genetic and molecular biology of autism spectrum disorder among Middle East population: a review | Wrong publication type |

| | | | |
|---|---|---|---|
| Nakhla (159) | 2021 | Assessment of 25 Hydroxy Cholecalceferol Level in Autistic Children; is there a role for it in Treatment of Autism Spectrum Disorder? | Wrong outcome |
| Shehata (160) | 2021 | Comparing levels of urinary phthalate metabolites in egyptian children with autism spectrum disorders and healthy control children: Referring to sources of phthalate exposure | Wrong outcome |
| Zaky (161) | 2021 | Neurodevelopmental Outcomes after Neonatal Mechanical Ventilation | Wrong publication type |
| Alotaibi (162) | 2021 | Sociodemographic, clinical characteristics, and service utilization of young children diagnosed with autism spectrum disorder at a research center in Saudi Arabia | Wrong comparison |
| Sadek (163) | 2021 | Clinical and laboratory characteristics of children with autism spectrum disorder at sohag university hospital | Wrong outcome |
| Alkhalidy (164) | 2021 | Nutritional Status of Pre-school Children and Determinant Factors of Autism: A Case-Control Study | Wrong outcome |
| Alrahili (165) | 2021 | The Association Between Screen Time Exposure and Autism Spectrum Disorder-Like Symptoms in Children | Wrong outcome |
| Zahra (166) | 2022 | Socio-economic Status in Egyptian Patients with Autism Spectrum Disorder. Does it affect Autism Severity? | Wrong comparison |
| Aloufi (167) | 2022 | Breastfeeding and Its Relation with Autism Spectrum Disorder in Children | Wrong outcome |
| Arafa (168) | 2022 | Maternal and neonatal risk factors for autism spectrum disorder: A case-control study from Egypt | Wrong outcome |
| Gholamalizadeh (169) | 2022 | The association of body mass index and dietary fat intake with autism in children: a case-control study | Retracted article |
| Kheirouri (170) | 2022 | Contribution of Excessive Gestational Weight Gain to the Increased Risk of Autism Spectrum Disorder Occurrence in Offspring | Wrong outcome |
| Sadiq (171) | 2022 | Toxoplasmosis is a risk factor in autism disease in Al-Diwaniyah governorate, Iraq | Wrong outcome |
| Slama (172) | 2022 | Risk factors in autism spectrum disorder: A Tunisian case-control study | Wrong outcome |
| Akbari (173) | 2022 | Association between angiotensin I converting enzyme gene polymorphisms and risk of autism in Iranian population | Wrong outcome |
| Lord (174) | 2022 | How matrix metalloproteinase (MMP)-9 (rs3918242) polymorphism affects MMP-9 serum concentration and associates with autism spectrum disorders: A case-control study in Iranian population | Wrong outcome |

| | | | |
|---|---|---|---|
| Hamed (175) | 2022 | Anti-ganglioside M1 autoantibodies in Egyptian children with autism: a cross-sectional comparative study | Wrong outcome |
| Hassan (176) | 2022 | Vitamin D3 status and polymorphisms of vitamin D receptor genes among cohort of Egyptian children with autism | Wrong outcome |
| Raouf (177) | 2022 | Association of immune abnormalities with symptom severity in Egyptian autistic children | Wrong outcome |
| Rezaei (178) | 2022 | A case-control study on the relationship between urine trace element levels and autism spectrum disorder among Iranian children | Wrong outcome |
| Al-Ali (179) | 2022 | The oxytocin receptor gene polymorphism rs2268491 and serum oxytocin alterations are indicative of autism spectrum disorder: A case-control paediatric study in Iraq with personalized medicine implications | Wrong outcome |
| Al-Dakroury (180) | 2022 | Autism in the Kingdom of Saudi Arabia: Current Situation and Future Perspectives for Services and Research | Wrong publication type |
| Aljumaili (181) | 2022 | Assessment of hair aluminium, cobalt, and mercury in a specimen of autistic Iraqi patients: Environmental risk factors of heavy metals in autism | Wrong outcome |
| Alenezi (182) | 2022 | Psychotropic Medications Use among Children with Autism in Saudi Arabia | Wrong comparison |
| Dehiol (183) | 2022 | Autism spectrum disorders and electronic screen devices exposure in Al-Nasiriya city 2019-2020 | Wrong outcome |
| Hajj (184) | 2022 | Pre-, Peri-, and Neonatal Factors Associated with Autism Spectrum Disorder: Results of a Lebanese Case-control Study | Wrong outcome |
| Aldera (185) | 2022 | Do Parental Comorbidities Affect the Severity of Autism Spectrum Disorder? | Wrong outcome |
| Sefrioui (186) | 2023 | Profile of autism spectrum disorders in Morocco: cross-sectional retrospective study of parents of children with autism | Wrong outcome |
| Alakhzami (187) | 2023 | Individuals with Autism Spectrum Disorders and Developmental Disorders in Oman: An Overview of Current Status | Wrong publication type |
| Alamoudi (188) | 2023 | Prenatal maternal stress and the severity of autism spectrum disorder: A cross-sectional study | Wrong comparison |
| Al-Awadi (189) | 2023 | Chromosomal aberration detection in Iraqi children with autism | Wrong outcome |
| Aljumaili (190) | 2023 | Determination of hair lead, iron, and cadmium in a sample of autistic Iraqi children: Environmental risk factors of heavy metals in autism | Wrong outcome |
| AlQahtani (191) | 2023 | Autism spectrum disorder: Where does the Gulf Region stand? An overview of ASD in the Arab Gulf Region: The UAE as a regional model | Wrong publication type |

| | | | |
|---|---|---|---|
| Alshaban (192) | 2023 | Consanguinity as a Risk Factor for Autism | Wrong outcome |
| Heidari (193) | 2023 | The Association Between Autism Spectrum Disorder and Attention Deficit Hyperactivity Disorder Symptoms in Medical Students | Wrong outcome |
| Jenabi (194) | 2023 | Autism Spectrum Disorders Registry in Hamadan, Iran: A Study Protocol | Wrong publication type |
| Jenabi (195) | 2023 | Is Breastfeeding Duration Associated with Risk of Developing ASD? | Full-text not retrieved |
| Kassab (196) | 2023 | Assessment of caries prevalence and related risk factors among a group of lebanese children with autism spectrum disorder: a case-control study | Wrong outcome |
| Meguid (197) | 2023 | Prevalence of autism spectrum disorder among children referred to special needs clinic in Giza | Wrong population |
| Nasir (198) | 2023 | Pediatricians' perspectives on childhood behavioral and mental health problems in Jordan | Wrong outcome |
| Saleh (199) | 2023 | Hair and Blood Levels of Aluminum, Cadmium, and Lead in Children with Autism from Egypt: Can Toxic Heavy Metals Increase the Risk of Autism? | Wrong outcome |
| Burhani (200) | 2023 | Perinatal Risk Factors in Children Diagnosed with Autism Spectrum Disorder in Dubai: A Case-Control Study | Wrong outcome |
| Al-Bishri (201) | 2023 | Glucose transporter 1 deficiency, AMP-activated protein kinase activation and immune dysregulation in autism spectrum disorder: Novel biomarker sources for clinical diagnosis | Wrong outcome |
| Hassan (202) | 2023 | Toxoplasmosis and cytomegalovirus infection and their role in Egyptian autistic children | Wrong outcome |
| Sadeghi (203) | 2023 | Associations between Symptom Severity of Autism Spectrum Disorder and Screen Time among Toddlers Aged 16 to 36 Months | Wrong outcome |
| Meimand (204) | 2023 | Burden of autism spectrum disorders in North Africa and Middle East from 1990 to 2019: A systematic analysis for the Global Burden of Disease Study 2019 | Wrong outcome |
| Abdi (205) | 2023 | Genomic architecture of autism spectrum disorder in Qatar: The BARAKA-Qatar Study | Wrong outcome |
| Moravej (206) | 2023 | Inborn Errors of Metabolism Associated With Autism Among Children: A Multicenter Study from Iran | Wrong comparison |
| Boujelben (207) | 2023 | Familial Autism Spectrum Disorder : A clinical study from South Tunisia | Wrong publication type |
| Ghahari (208) | 2023 | Prenatal exposure to ambient air pollution and autism spectrum disorders: Results from a family-based case-control study | Wrong outcome |

| | | | |
|---|---|---|---|
| Zebbiche (209) | 2024 | Trace element levels and autism spectrum disorder in a sample of Algerian children: A case-control study investigation | Wrong outcome |

**Assessment of the Quality of Included Studies**

Supplementary Table S9. Scores for the assessment of the quality of included cross sectional studies using the adapted Newcastle-Ottawa quality assessment scale for cross sectional studies.

| Study | A (max 3) | B (max 2) | C (max 2) | D (max 2) | E (max 1) | F (max 1) | G (max 3) | H (max 1) | Total |
|---|---|---|---|---|---|---|---|---|---|
| Al-Farsi, 2011 | 3 | 2 | 2 | 2 | 0 | 0 | 3 | 1 | 13 |
| Samadi, 2012 | 3 | 2 | 2 | 2 | 1 | 1 | 3 | 1 | 15 |
| Zeglam, 2012 | 3 | 2 | 2 | 2 | 1 | 0 | 3 | 1 | 14 |
| Goodarzi, 2019 | 2 | 0 | 0 | 2 | 0 | 0 | 3 | 1 | 8 |
| Al-Mamri, 2019 | 2 | 1 | 0 | 2 | 0 | 0 | 2 | 1 | 8 |
| Mohammadi, 2019 | 3 | 2 | 0 | 2 | 1 | 0 | 3 | 1 | 12 |
| Yousef, 2021 | 2 | 2 | 0 | 2 | 0 | 1 | 3 | 1 | 11 |
| Sabbagh, 2021 | 2 | 1 | 1 | 2 | 0 | 0 | 2 | 1 | 9 |
| AlBatti, 2022 | 2 | 2 | 2 | 2 | 1 | 1 | 1 | 1 | 12 |
| Samadi,2015 | 2 | 2 | 2 | 2 | 0 | 0 | 3 | 0 | 11 |
| Chaaya, 2016 | 2 | 2 | 2 | 0 | 0 | 0 | 0 | 1 | 7 |
| Eapen, 2007 | 3 | 1 | 1 | 2 | 0 | 0 | 3 | 1 | 11 |
| Manzouri, 2019 | 1 | 2 | 0 | 2 | 0 | 0 | 3 | 1 | 9 |
| Alshaban, 2019 | 3 | 2 | 1 | 2 | 0 | 0 | 3 | 1 | 12 |

Supplementary Table S10. Scores for the assessment of the quality of included case control studies using the Newcastle-Ottawa quality assessment scale for case control studies.

| Study | A (max 2) | B (max 1) | C (max 2) | D (max 1) | E (max 1) | F (max 1) | G (max 4) | H (max 1) | I (max 2) | Total |
|---|---|---|---|---|---|---|---|---|---|---|
| Al-Ansari, 2013 | 1 | 1 | 1 | 1 | 0 | 1 | 4 | 1 | 0 | 10 |
| Sasanfar, 2010 | 2 | 1 | 2 | 1 | 1 | 0 | 3 | 1 | 2 | 13 |

**Supplementary Tables**

Supplementary Table S11. Summary of Egger's *p* Values for the analyzed risk factors.

| **Prevalence** | **Egger's *p* Value** |
|---|---|
| Prevalence in MENA | 0.150 |
| Prevalence in Iran | 0.003 |

**Supplementary Figures**

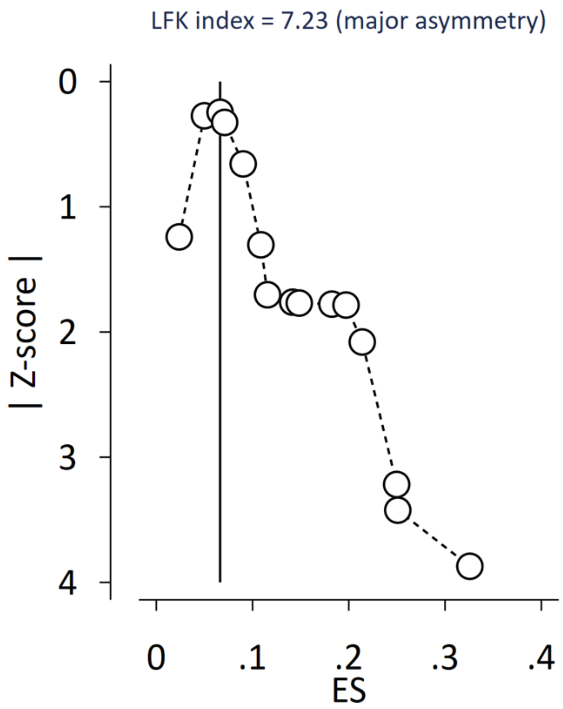

Supplementary Figure S1. Doi Plot and LFK index for the prevalence of ASD in MENA region.

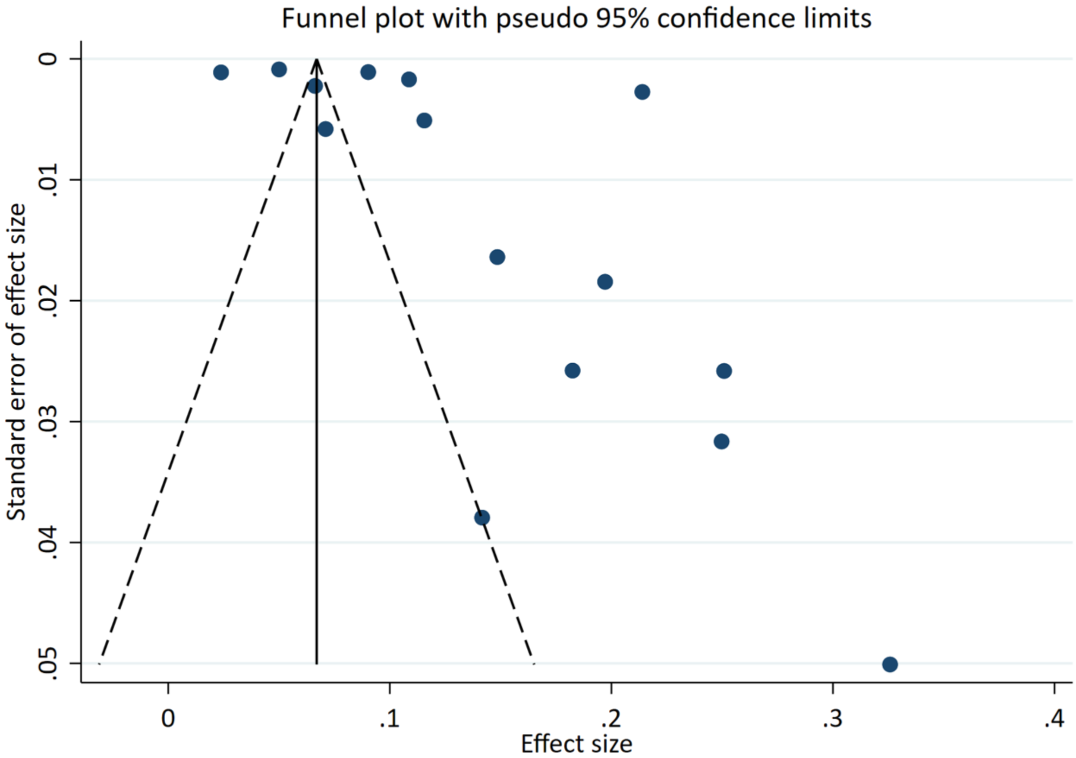

Supplementary Figure S2. Funnel Plot for the prevalence of ASD in MENA region.

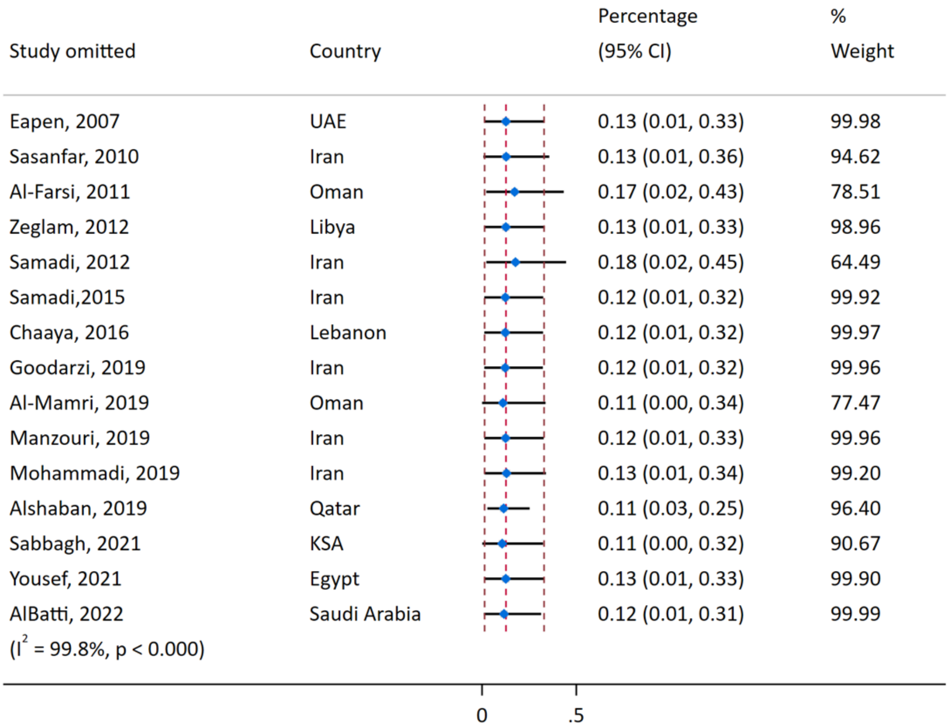

Supplementary Figure S3. Leave-one-out analysis for the prevalence of ASD in MENA region.

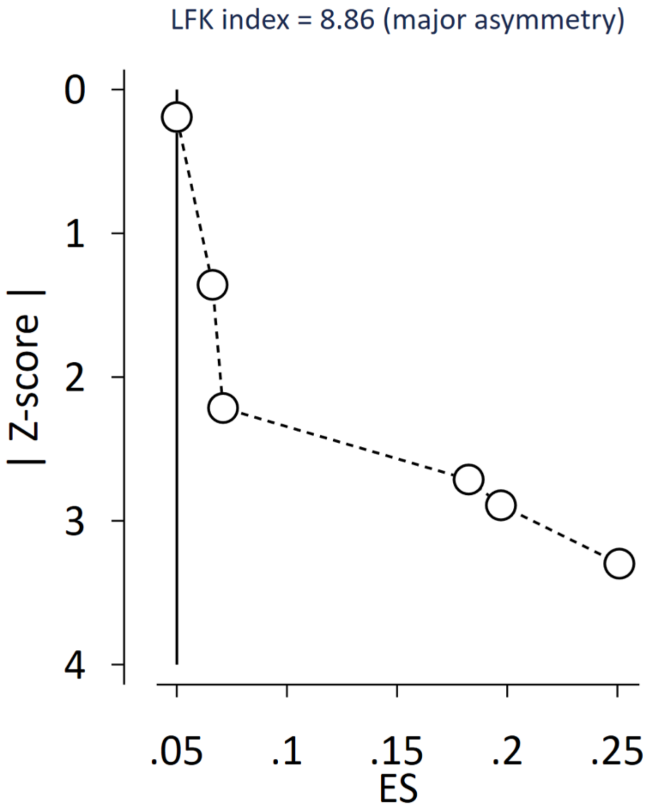

Supplementary Figure S4. Doi Plot and LFK index for the prevalence of ASD in Iran.

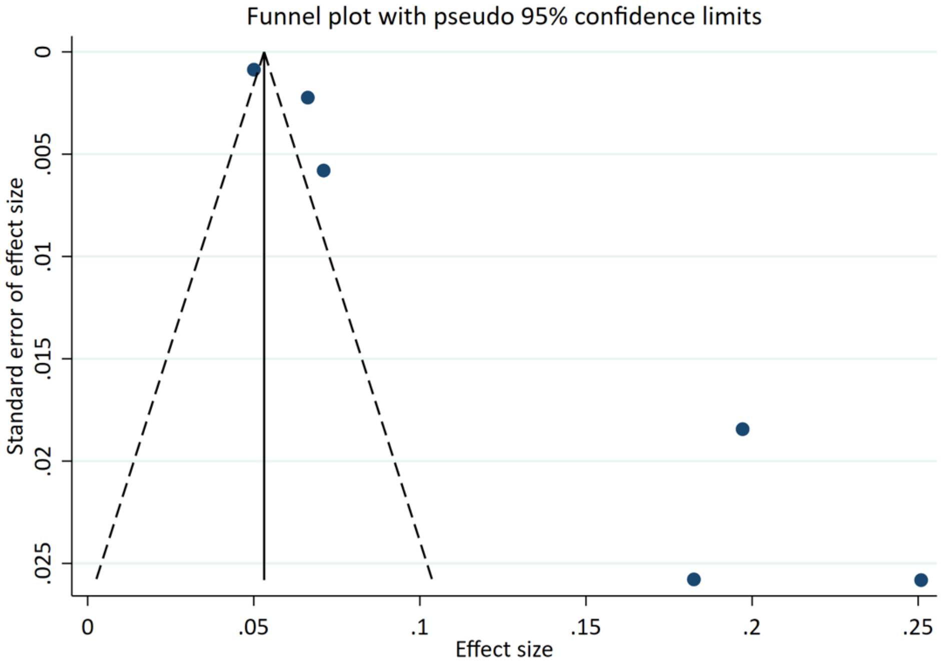

Supplementary Figure S5. Funnel Plot for the prevalence of ASD in Iran.

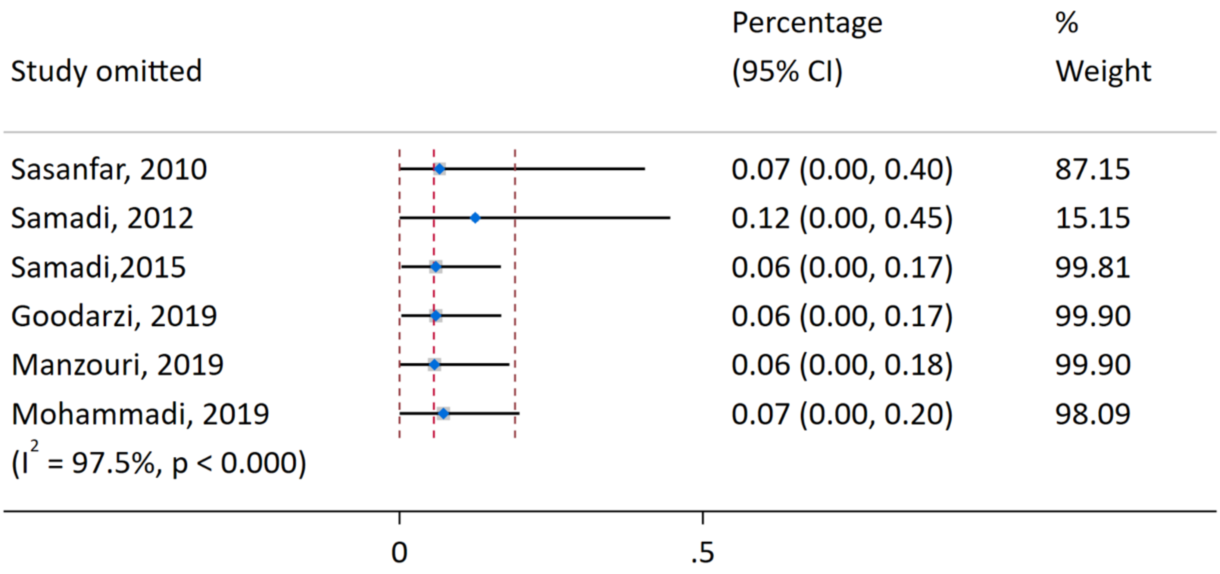

Supplementary Figure S6. Leave-one-out analysis for the prevalence of ASD in Iran.

### References

1. Eapen V, Ghubash R, Zoubeidi T, Yunis F, Aithala G, Sabri S. Epidemiological study of developmental and behavioural disorders in three year old children: A UAE study. Emirates Medical Journal. 2003;21(3):237-42.
2. Al-Ayadhi LY. Heavy metals and trace elements in hair samples of autistic children in central Saudi Arabia. Neurosciences. 2005;10(3):213-8.
3. Al-Ayadhi LY. Pro-inflammatory cytokines in autistic children in central Saudi Arabia. Neurosciences. 2005;10(2):155-8.
4. Al-Ayadhi LY. Autoimmune connection of autism in Central Saudi Arabia. Neurosciences. 2005;10(4):265-7.
5. Al-Ayadhi LY. Altered oxytocin and vasopressin levels in autistic children in Central Saudi Arabia. Neurosciences. 2005;10(1):47-50.
6. Mostafa GA, El-Sayed ZA, Manal MMA, El-Sayed MF. Serum anti-myelin - Associated glycoprotein antibodies in Egyptian Autistic children. Journal of Child Neurology. 2008;23(12):1413-8.
7. al-Salehi SM, al-Hifthy EH, Ghaziuddin M. Autism in Saudi Arabia: PresentatioN, Clinical Correlates and Comorbidity. Transcultural Psychiatry. 2009;46(2):340-7.
8. Walsh CA, Yu T, Chahrour M, Schubert C, Hill S. High-throughput dna sequencing in autism spectrum disorders (ASD). Neuropsychopharmacology. 2010;35:S5.
9. Meguid NA, Hashish AF, Anwar M, Sidhom G. Reduced serum levels of 25-hydroxy and 1,25-dihydroxy vitamin D in Egyptian children with autism. Journal of Alternative and Complementary Medicine. 2010;16(6):641-5.
10. El-Ansary A, Al-Daihan S, Al-Dbass A, Al-Ayadhi L. Measurement of selected ions related to oxidative stress and energy metabolism in Saudi autistic children. Clinical Biochemistry. 2010;43(1):63-70.
11. Al-Farsi YM, Al-Sharbati MM, Waly MI, Al-Farsi OA, Al Shafaee MA, Deth RC. Malnutrition among preschool-aged autistic children in Oman. Research in Autism Spectrum Disorders. 2011;5(4):1549-52.
12. Al-Yafee YA, Al- Ayadhi LY, Haq SH, El-Ansary AK. Novel metabolic biomarkers related to sulfur-dependent detoxification pathways in autistic patients of Saudi Arabia. BMC Neurology. 2011;11.

13. El-Ansary AK, Ben Bacha AG, Al-Ayadhi LY. Proinflammatory and proapoptotic markers in relation to mono and di-cations in plasma of autistic patients from Saudi Arabia. Journal of Neuroinflammation. 2011;8.
14. Hussein H, Taha GRA, Almanasef A. Characteristics of autism spectrum disorders in a sample of egyptian and saudi patients: Transcultural cross sectional study. Child and Adolescent Psychiatry and Mental Health. 2011;5.
15. Meguid NA, Dardir AA, Abdel-Raouf ER, Hashish A. Evaluation of oxidative stress in autism: Defective antioxidant enzymes and increased lipid peroxidation. Biological Trace Element Research. 2011;143(1):58-65.
16. Mohareri F, Soltanifar A. Attention deficit hyperactivity symptoms in children with autistic spectrum disorder. European Psychiatry. 2011;26.
17. Abd Elhameed MA, Abd Elbaky AEO, Kamel EA. A controlled study of the risk factors and clinical picture of children with Autism in an Egyptian sample. Egyptian Journal of Neurology, Psychiatry and Neurosurgery. 2011;48(3):271-6.
18. El-Baz F, Ismael NA, El-Din SMN. Risk factors for autism: An Egyptian study. Egyptian Journal of Medical Human Genetics. 2011;12(1):31-8.
19. Waly MI, Al-Farsi YM, Al-Sharbati MM, Ali A, Al-Khalili MM, Essa MM, et al. Low serum levels of glutathione, homocysteine and total antioxidant capacity are associated with increased risk of autism in Oman. FASEB Journal. 2011;25.
20. Amr M, Raddad D, El-Mehesh F, Mahmoud EH, El-Gilany AH. Sex differences in Arab children with Autism spectrum disorders. Research in Autism Spectrum Disorders. 2011;5(4):1343-50.
21. Mohammadi MR, Salmanian M, Akhondzadeh S. Autism spectrum disorders in Iran. Iranian Journal of Child Neurology. 2011;5(4):1-9.
22. Masri A, Hamamy H, Khreisat A. Profile of developmental delay in children under five years of age in a highly consanguineous community: A hospital-based study - Jordan. Brain and Development. 2011;33(10):810-5.
23. Gebril OH, Meguid NA. HFE gene polymorphisms and the risk for autism in Egyptian children and impact on the effect of oxidative stress. Disease Markers. 2011;31(5):289-94.
24. El-Ansary AK, Ben Bacha AG, Al- Ayahdi LY. Impaired plasma phospholipids and relative amounts of essential polyunsaturated fatty acids in autistic patients from Saudi Arabia. Lipids in Health and Disease. 2011;10.

25. Zeglam AM, Maouna A. Is there a need for a focused health care service for children with autistic spectrum disorders? A keyhole look at this problem in Tripoli, Libya. Autism. 2012;16(4):337-9.
26. El-Ansary A, Al-Ayadhi L. Lipid mediators in plasma of autism spectrum disorders. Lipids in Health and Disease. 2012;11(1).
27. El-Ansary A, Bacha AB, Al-Ayahdi L. Relationship between chronic lead toxicity and plasma neurotransmitters in autistic patients from Saudi Arabia. Brain Injury. 2012;26(4):310.
28. Essa MM, Guillemin GJ, Waly MI, Al-Sharbati MM, Al-Farsi YM, Hakkim FL, et al. Increased markers of oxidative stress in autistic children of the Sultanate of Oman. Biological Trace Element Research. 2012;147(1):25-7.
29. Elshahawi HH. Possible association of certain HLA-DRB1 alleles with autism in Egyptian children: Relation to family history of autoimmunity. Neuropsychiatrie de l'Enfance et de l'Adolescence. 2012;60(5):S220.
30. Zakareia FA, Al-Ayadhi LY, Al-Drees AMA. Study of dual angiogenic/neurogenic growth factors among Saudi autistic children and their correlation with the severity of this disorder. Neurosciences. 2012;17(3):213-8.
31. Amr M, Raddad D, El-Mehesh F, Bakr A, Sallam K, Amin T. Comorbid psychiatric disorders in Arab children with Autism spectrum disorders. Research in Autism Spectrum Disorders. 2012;6(1):240-8.
32. Al-Sharbati M, Al-Farsi Y, Al-Sharbati Z, Ouhtit A, Waly M, Al-Khaduri M, et al. Autistic Spectrum Disorder (ASD) Among Omani Children Below 6 Years: A five-year retrospective descriptive study. 2012 International Meeting for Autism Research2012.
33. Al-Farsi YM, Waly MI, Deth RC, Al-Sharbati MM, Al-Shafaee M, Al-Farsi O, et al. Impact of nutrition on serum levels of docosahexaenoic acid among Omani children with autism. Nutrition. 2013;29(9):1142-6.
34. Al-Farsi YM, Waly MI, Al-Sharbati MM, Al-Shafaee MA, Al-Farsi OA, Al-Khaduri MM, et al. Levels of heavy metals and essential minerals in hair samples of children with autism in Oman: A case-control study. Biological Trace Element Research. 2013;151(2):181-6.
35. Al-Rubaye FG, Morad TS. Purine metabolism and oxidative stress in children with autistic spectrum disorders. Journal of Experimental and Integrative Medicine. 2013;3(4):293-7.
36. Zakareia FA, Al-Ayadhi LY. Evaluation of plasma soluble fatty acid synthase levels among Saudi autistic children: Relation to disease severity. Neurosciences. 2013;18(3):242-7.

37. Al-Farsi YM, Al-Khaduri M, Al-Sumri H, Al-Farsi O, Al-Sharbati M, Waly M, et al. Association of gestational diabetes mellitus with occurrence of Autism: A cohort study. FASEB Journal. 2013;27.

38. Bashir S, Halepoto DM, Al-Ayadhi L. Serum level of desert hedgehog protein in autism spectrum disorder: Preliminary results. Medical Principles and Practice. 2013;23(1):14-7.

39. Hamadé A, Salameh P, Medlej-Hashim M, Hajj-Moussa E, Saadallah-Zeidan N, Rizk F. Autism in children and correlates in lebanon: A pilot case-control study. Journal of Research in Health Sciences. 2013;13(2):119-24.

40. Hashim H, Abdelrahman H, Mohammed D, Karam R. Association between plasma levels of transforming growth factor-β1, IL-23 and IL-17 and the severity of autism in Egyptian children. Research in Autism Spectrum Disorders. 2013;7(1):199-204.

41. Mousavizadeh K, Askari M, Arian H, Gorjipour F, Nikpour AR, Tavafjadid M, et al. Association of human mtDNA mutations with autism in Iranian patients. Journal of Research in Medical Sciences. 2013;18(10):926.

42. Kaddah FAA, Nassar JF, Ghandour HH, El-Farghali OG. Screening for autism in low-birth-weight Egyptian toddlers. Egyptian Journal of Otolaryngology. 2013;29(1):38-45.

43. Al-Hakbany M, Awadallah S, Al-Ayadhi L. The Relationship of HLA Class I and II Alleles and Haplotypes with Autism: A Case Control Study. Autism Res Treat. 2014;2014:242048.

44. Ranjbar A, Rashedi V, Rezaei M. Comparison of urinary oxidative biomarkers in Iranian children with autism. Research in Developmental Disabilities. 2014;35(11):2751-5.

45. Hodgson NW, Waly MI, Al-Farsi YM, Al-Sharbati MM, Al-Farsi O, Ali A, et al. Decreased glutathione and elevated hair mercury levels are associated with nutritional deficiency-based autism in Oman. Experimental Biology and Medicine. 2014;239(6):697-706.

46. Yassa HA. Autism: A form of lead and mercury toxicity. Environmental Toxicology and Pharmacology. 2014;38(3):1016-24.

47. Alabdali A, Al-Ayadhi L, El-Ansary A. Association of social and cognitive impairment and biomarkers in autism spectrum disorders. Journal of Neuroinflammation. 2014;11.

48. El-Ansary A, Hassan SA, Anwar M, Shmais GA, Bhat RS, Hashish A, et al. Role of amino acids in the pathophysiology of autism spectrum disorders in Saudi and Egyptian population samples. Journal of Pediatric Neurology. 2014;12(4):171-81.

49. Shawky RM, El-baz F, Kamal TM, Elhossiny RM, Ahmed MA, El Nady GH. Study of genotype-phenotype correlation of methylene tetrahydrofolate reductase (MTHFR) gene polymorphisms in a sample of Egyptian autistic children. Egyptian Journal of Medical Human Genetics. 2014;15(4):335-41.
50. Afsharpaiman S, Skandari A, Jahromi MZ, Radfar S, Shirbazoo S, Amirsalari S, et al. An assessment of toxoplasmosis antibodies seropositivity in children suffering autism. Tehran University Medical Journal. 2014;72(2):106-12.
51. Mohamed FEB, Zaky EA, El-Sayed AB, Elhossieny RM, Zahra SS, Salah Eldin W, et al. Assessment of Hair Aluminum, Lead, and Mercury in a Sample of Autistic Egyptian Children: Environmental Risk Factors of Heavy Metals in Autism. Behavioural Neurology. 2015;2015.
52. Halepoto DM, Bashir S, Zeina R, Al-Ayadhi LY. Correlation Between Hedgehog (Hh) Protein Family and Brain-Derived Neurotrophic Factor (BDNF) in Autism Spectrum Disorder (ASD). Journal of the College of Physicians and Surgeons Pakistan. 2015;25(12):882-5.
53. Fanid LM, Shahrokhi H, Adampourezare M, Feizi MAH, Bonyadi M, Eslami A. Association between common single- nucleotide polymorphism of reelin gene, rs736707 (C/T) with autism spectrum disorder in Iranian-Azeri patients. International Journal of Pediatrics. 2015;3(6):1065-71.
54. Fernell E, Bejerot S, Westerlund J, Miniscalco C, Simila H, Eyles D, et al. Autism spectrum disorder and low vitamin D at birth: A sibling control study. Molecular Autism. 2015;6(1).
55. Slama H, Brahim T, Attia M, Gadour N, Missaoui S. Family history of psychiatric disorder and autism spectrum disorders: A study about 790 cases. European Child and Adolescent Psychiatry. 2015;24(1):S199.
56. Meguid N, Khalil R, Gebril O, El-Fishawy P. Evaluation of MTHFR genetic polymorphism as a risk factor in Egyptian autistic children and mothers. African Journal of Psychiatry (South Africa). 2015;18(1).
57. Mousavi SM, Kazemi M, Salehi M, Kamali E, Karimi P, Karahmadi M. RoRa gene contribution to autism; another epigenetic layer on autism complexity. Genetics in the Third Millennium. 2015;12(4):3726-35.
58. Ouhtit A, Al-Farsi Y, Al-Sharbati M, Waly M, Gupta I, Al-Farsi O, et al. Underlying factors behind the low prevalence of autism spectrum disorders in oman sociocultural perspective. Sultan Qaboos University Medical Journal. 2015;15(2):e213-e7.

59. Yavarna T, Al-Dewik N, Al-Mureikhi M, Ali R, Al-Mesaifri F, Mahmoud L, et al. High diagnostic yield of clinical exome sequencing in Middle Eastern patients with Mendelian disorders. Human Genetics. 2015;134(9):967-80.
60. Khakzad MR, Salari F, Javanbakht M, Hojati M, Varasteh A, Sankian M, et al. Transforming growth factor beta 1 869T/C and 915G/C polymorphisms and risk of autism spectrum disorders. Rep Biochem Mol Biol. 2015;3(2):82-8.
61. Afrazeh M, Saedisar S, Khakzad MR, Hojati M. Measurement of Serum Superoxide Dismutase and Its Relevance to Disease Intensity Autistic Children. Maedica (Bucur). 2015;10(4):315-8.
62. Saad K, Elserogy Y, Abdel Rahman AA, Al-Atram AA, Mohamad IL, ElMelegy TT, et al. ADHD, autism and neuroradiological complications among phenylketonuric children in Upper Egypt. Acta Neurol Belg. 2015;115(4):657-63.
63. Al-Sharbati MM, Al-Farsi YM, Al-Sharbati ZM, Al-Sulaimani F, Ouhtit A, Al-Adawi S. Profile of mental and behavioral disorders among preschoolers in a tertiary care hospital in Oman: A retrospective study. Oman Medical Journal. 2016;31(5):357-64.
64. Fahmy SF, Sabri NA, El Hamamsy MH, El Sawi M, Zaki OK. Vitamin D intake and sun exposure in autistic children. International Journal of Pharmaceutical Sciences and Research. 2016;7(3):1043-9.
65. Haghiri R, Mashayekhi F, Bidabadi E, Salehi Z. Analysis of methionine synthase (rs1805087) gene polymorphism in autism patients in northern Iran. Acta Neurobiologiae Experimentalis. 2016;76(4):318-23.
66. El-Ansary A. Data of multiple regressions analysis between selected biomarkers related to glutamate excitotoxicity and oxidative stress in Saudi autistic patients. Data in Brief. 2016;7:111-6.
67. Environmental toxic pollutant and trace elements in Egyptian children with autism. Developmental Medicine & Child Neurology. 2016;58:10-1.
68. Dinkler L, Lundstrom S, Gajwani R, Lichtenstein P, Gillberg C, Minnis H. Maltreatment-associated neurodevelopmental problems: Environment and genetics. Behavior Genetics. 2016;46(6):779-80.
69. Elhawary N, Tayeb M, Bogari N, Qotb N. Vulnerability of genetic variants to the risk of autism among Saudi children. Human Genomics. 2016;10.
70. Fanid LM, Feizi MAH, Zare MA, Shahrokhi H. An Association Analysis of Reelin Gene (RELN) exon 22 (G/C), Rs.362691, polymorphism with autism spectrum disorder among Iranian-Azeri population. International Journal of Pediatrics. 2016;4(7):2027-33.

71. Khaled EM, Meguid NA, Bjørklund G, Gouda A, Bahary MH, Hashish A, et al. Altered urinary porphyrins and mercury exposure as biomarkers for autism severity in Egyptian children with autism spectrum disorder. Metabolic Brain Disease. 2016;31(6):1419-26.

72. Meguid NA, Azab SN, Saber AS, Mostafa HA, El-Bana MA, Hashish A, et al. Impact of oxidative stress on autism spectrum disorder behaviors in children with autism. International Journal of Pharmaceutical and Clinical Research. 2016;8(3):193-7.

73. Mohammed HS, Wahass SH, Mahmoud AA. Incidence of autism in high risk neonatal follow up. Neurosciences. 2016;21(1):43-6.

74. Alhowikan AM, Al-Ayadhi L, Halepoto DM. Secretagogin (SCGN) plasma levels and their association with cognitive and social behavior in children with autism spectrum disorder (ASD). Journal of the College of Physicians and Surgeons Pakistan. 2017;27(4):222-6.

75. Alshaban F, Aldosari M, Sayed ZE, Tolefat M, Shammari SEHHA, Ghazal I, et al. Autism spectrum disorder in Qatar: Profiles and correlates of a large clinical sample. Autism and Developmental Language Impairments. 2017;2.

76. Bener A, Khattab A, Bhugra D, Hoffmann G. Iron and Vitamin D levels among autism spectrum disorders children. Annals of African Medicine. 2017;16(4):186-91.

77. Desoky T, Hassan MH, Fayed HM, Sakhr HM. Biochemical assessments of thyroid profile, serum 25-hydroxycholecalciferol and cluster of differentiation 5 expression levels among children with autism. Neuropsychiatric Disease and Treatment. 2017;13:2397-403.

78. Ashaat EA, Taman KH, Kholoussi N, El Ruby MO, Zaki ME, El Wakeel MA, et al. Altered adaptive cellular immune function in a group of Egyptian children with autism. Journal of Clinical and Diagnostic Research. 2017;11(10):SC14-SC7.

79. Firouzabadi SG, Kariminejad R, Vameghi R, Darvish H, Ghaedi H, Banihashemi S, et al. Copy Number Variants in Patients with Autism and Additional Clinical Features: Report of VIPR2 Duplication and a Novel Microduplication Syndrome. Molecular Neurobiology. 2017;54(9):7019-27.

80. Khaniani MS, Yeganeh FA, Amiri S, Derakhshan SM. Autistic Phenotype of Permutation and Intermediate Alleles of FMR1 Gene. Iranian Journal of Pediatrics. 2017;27(4):1-5.

81. Kourtian S, Soueid J, Makhoul NJ, Guisso DR, Chahrour M, Boustany RMN. Candidate Genes for Inherited Autism Susceptibility in the Lebanese Population. Scientific Reports. 2017;7.

82. Zeglam A, Al-Ogab M, Tubbal AL. Epidemiology of autism in Libya: Uncovering of the first autism findings. Journal of the Neurological Sciences. 2017;381:931.

83. Dinkler L, Lundström S, Gajwani R, Lichtenstein P, Gillberg C, Minnis H. Maltreatment-associated neurodevelopmental disorders: a co-twin control analysis. Journal of Child Psychology and Psychiatry and Allied Disciplines. 2017;58(6):691-701.

84. Ajabi S, Mashayekhi F, Bidabadi E. A study of MTRR 66A>G gene polymorphism in patients with autism from northern Iran. Neurology Asia. 2017;22(1):59-64.

85. Hosseinpour M, Mashayekhi F, Bidabadi E, Salehi Z. Neuropilin-2 rs849563 gene variations and susceptibility to autism in Iranian population: A case-control study. Metabolic Brain Disease. 2017;32(5):1471-4.

86. Hamedani SY, Gharesouran J, Noroozi R, Sayad A, Omrani MD, Mir A, et al. Ras-like without CAAX 2 (RIT2): a susceptibility gene for autism spectrum disorder. Metabolic Brain Disease. 2017;32(3):751-5.

87. El-Ansary A, Bjørklund G, Tinkov AA, Skalny AV, Al Dera H. Relationship between selenium, lead, and mercury in red blood cells of Saudi autistic children. Metabolic Brain Disease. 2017;32(4):1073-80.

88. Noroozi R, Ghafouri-Fard S, Omrani MD, Habibi M, Sayad A, Taheri M. Association study of the vesicular monoamine transporter 1 (VMAT1) gene with autism in an Iranian population. Gene. 2017;625:10-4.

89. Safari MR, Omrani MD, Noroozi R, Sayad A, Sarrafzadeh S, Komaki A, et al. Synaptosome-Associated Protein 25 (SNAP25) Gene Association Analysis Revealed Risk Variants for ASD, in Iranian Population. Journal of Molecular Neuroscience. 2017;61(3):305-11.

90. Sayad A, Noroozi R, Omrani MD, Taheri M, Ghafouri-Fard S. Retinoic acid-related orphan receptor alpha (RORA) variants are associated with autism spectrum disorder. Metabolic Brain Disease. 2017;32(5):1595-601.

91. Zare S, Mashayekhi F, Bidabadi E. The association of CNTNAP2 rs7794745 gene polymorphism and autism in Iranian population. Journal of Clinical Neuroscience. 2017;39:189-92.

92. Zeglam A, Alogab M, Tubbal A. Early TV viewing and autistic spectrum disorder; A plausible hypothesis that should not be dismissed “Libyan Viewpoint”. Journal of the Neurological Sciences. 2017;381:931.

93. Abdulamir HA, Abdulhussain HA, Al-Awadi SJA, Abdul-Rasheed OF, Abdulghani EA. Acetylserotonin O-Methyltransferase (ASMT)/rs4446909 Polymorphism in Iraqi Autistic Children. Clinical Chemistry. 2018;64:S234.

94. Kadhim SJ, Abbas AAH, Abdul-Jabbar OK. Sero-positivity rate of rubella antibodies in Iraqi autistic children. Journal of Pharmaceutical Sciences and Research. 2018;10(5):1142-4.

95. Belkady B, Elkhattabi L, Elkarhat Z, Zarouf L, Razoki L, Aboulfaraj J, et al. Chromosomal Abnormalities in Patients with Intellectual Disability: A 21-Year Retrospective Study. Hum Hered. 2018;83(5):274-82.

96. Delgado AM, Pandey MK, DeGrado TR, Howie FR, Huebner AR, Mehta SQ. 5.28 Role of Metal Ion Dyshomeostasis in ASD: Evaluation of Copper, Zinc, and Selenium Levels in the North American ASD Population. Journal of the American Academy of Child and Adolescent Psychiatry. 2018;57(10):S235.

97. Alshiban A, Alomar M, Al-Ayadhi L. Risk factors for Autism Spectrum Disorder (ASD) in Saudi Arabia. European Psychiatry. 2018;48:S371.

98. Mousavi SM, Kamali E, Fatahi F, Babaie H, Salehi M. Autism and probable prerequisites: Severe and scheduled prenatal stresses at spotlight. Iranian Journal of Public Health. 2018;47(9):1387-95.

99. Yousefian F, Mahvi AH, Yunesian M, Hassanvand MS, Kashani H, Amini H. Long-term exposure to ambient air pollution and autism spectrum disorder in children: A case-control study in Tehran, Iran. Science of the Total Environment. 2018;643:1216-22.

100. John J, Sala R. Is the prevalence of autism spectrum disorder decreased in black and ethnic children and adolescents? Archives of Disease in Childhood. 2018;103:A17-A8.

101. Abdulamir HA, Abdul-Rasheed OF, Abdulghani EA. Serotonin and serotonin transporter levels in autistic children. Saudi Medical Journal. 2018;39(5):487-94.

102. Meguid NA, Nashaat NH, Hashem HS, Khalil MM. Frequency of risk factors and coexisting abnormalities in a population of Egyptian children with autism spectrum disorder. Asian Journal of Psychiatry. 2018;32:54-8.

103. Olusanya BO, Davis AC, Wertlieb D, Boo NY, Nair MKC, Halpern R, et al. Developmental disabilities among children younger than 5 years in 195 countries and territories, 1990–2016: a systematic analysis for the Global Burden of Disease Study 2016. The Lancet Global Health. 2018;6(10):e1100-e21.

104. Altamimi M. Could Autism Be Associated With Nutritional Status in the Palestinian population? The Outcomes of the Palestinian Micronutrient Survey. Nutrition and Metabolic Insights. 2018;11.

105. Noroozi R, Taheri M, Movafagh A, Ghafouri-Fard S, Sayad A, Mirfakhraie R, et al. Association analysis of the GABRB3 promoter variant and susceptibility to autism spectrum disorder. Basal Ganglia. 2018;11:4-7.

106. Qasem H, Al-Ayadhi L, Bjørklund G, Chirumbolo S, El-Ansary A. Impaired lipid metabolism markers to assess the risk of neuroinflammation in autism spectrum disorder. Metabolic Brain Disease. 2018;33(4):1141-53.

107. Arastoo AA, Khojastehkia H, Rahimi Z, Khafaie MA, Hosseini SA, Mansoori SM, et al. Evaluation of serum 25-Hydroxy vitamin D levels in children with autism Spectrum disorder. Italian Journal of Pediatrics. 2018;44(1).

108. Jabbar ARAA, Jebor MA. Study of polymorphism in methionine synthase gene by RFLP-PCR in middle euphrates region of Iraq. Journal of Pharmaceutical Sciences and Research. 2018;10(12):3219-22.

109. Eftekharian MM, Noroozi R, Omrani MD, Arsang-Jang S, Komaki A, Taheri M, et al. Expression Analysis of Protein Inhibitor of Activated STAT (PIAS) Genes in Autistic Patients. Advances in Neuroimmune Biology. 2018;7(2):129-34.

110. Sayad A, Akbari MT, Noroozi R, Omrani MD, Inoko H, Taheri M, et al. Association of HLA alleles with autism. Neuropsychiatric Disease and Treatment. 2018;14:3259-65.

111. Guisso DR, Saadeh FS, Saab D, El Deek J, Chamseddine S, El Hassan HA, et al. Association of Autism with Maternal Infections, Perinatal and Other Risk Factors: A Case-Control Study. Journal of Autism and Developmental Disorders. 2018;48(6):2010-21.

112. Oommen A, AlOmar RS, Osman AA, Aljofi HE. Role of environmental factors in autism spectrum disorders in Saudi children aged 3-10 years in the Northern and Eastern regions of Saudi Arabia. Neurosciences (Riyadh, Saudi Arabia). 2018;23(4):286-91.

113. Alawad ZM, Al-Jobouri SM, Majid AY. Lead among children with autism in Iraq. Is it a potential factor?: Lead level in Iraqi children with autism. Journal of Clinical and Analytical Medicine. 2019;10(2):215-9.

114. Alzghoul L, Abdelhamid SS, Yanis AH, Qwaider YZ, Aldahabi M, Albdour SA. The association between levels of inflammatory markers in autistic children compared to their unaffected siblings and unrelated healthy controls. Turkish Journal of Medical Sciences. 2019;49(4):1047-53.

115. Elsayed S, Omar T, El Bardeny M, El-Latif SA, Ibrahim S. Study of autistic features among children and adolescents with congenital adrenal hyperplasia. Hormone Research in Paediatrics. 2019;91:481.

116. Jabbar ARAA, Jebor MA. Evaluation the Relationship Between DRD1 RS4532 Gene Polymorphisms and Autism by RFLP-PCR in Middle Euphrates Region of Iraq. Biochemical and Cellular Archives. 2019;19(1):57-61.

117. Alhader AA, Alfaqih MM, Nazzal MS. 1.11 THE POTENTIAL INTERACTIVE ROLE OF LEPTIN WITH OTHER HORMONES IN THE PATHOPHYSIOLOGY OF ASD IN JORDANIAN MALE CHILDREN. Journal of the American Academy of Child and Adolescent Psychiatry. 2019;58(10):S150.

118. Arab AH, Elhawary NA. Methylenetetrahydrofolate reductase gene variants confer potential vulnerability to autism spectrum disorder in a Saudi community. Neuropsychiatric Disease and Treatment. 2019;15:3569-81.

119. El Khatib R, Hajj A, Hallit S, Itani T, Karam Sarkis D. Gastrointestinal symptoms in children with autism spectrum disorders and correlation with autism functionality and severity: a case-control study. European Psychiatry. 2019;56:S33.

120. Ismail S, Senna AA, Behiry EG, Ashaat EA, Zaki MS, Ashaat NA, et al. Study of C677T variant of methylene tetrahydrofolate reductase gene in autistic spectrum disorder Egyptian children. American Journal of Medical Genetics, Part B: Neuropsychiatric Genetics. 2019;180(5):305-9.

121. Sayad A, Ghafouri-Fard S, Noroozi R, Omrani MD, Ganji M, Dastmalchi R, et al. Association study of sequence variants in voltage-gated Ca2+ channel subunit alpha-1C and autism spectrum disorders. Reports of Biochemistry and Molecular Biology. 2019;8(1).

122. Bordeleau M, Luheshi G, Tremblay MÈ. Microglia as a potential link between pathological myelination and stereotypic behavior after exposure to maternal high-fat diet. IBRO Reports. 2019;6:S515-S6.

123. Gleeson JG. The genetic and molecular basis of neurodevelopmental disorders. Medizinische Genetik. 2019;31(1):72.

124. Hasan AA. Assessment of children upon suffering from withdrawals signs in Baghdad city, Iraq. Indian Journal of Forensic Medicine and Toxicology. 2019;13(4).

125. Malek A, Farhang S, Amiri S, Abdi S, Rezaih AR, Asadian M. Risk factors for autistic disorder: A case-control study. Iranian Journal of Pediatrics. 2019;29(3).

126. Ben Sadek AT, Bakarman MA. Factors associated with autism in Saudi Arabia: A case-control study. Pakistan Paediatric Journal. 2019;43(1):30-5.

127. Al-Zalabani AH, Al-Jabree AH, Zeidan ZA. Is cesarean section delivery associated with autism spectrum disorder? Neurosciences. 2019;24(1):11-5.
128. Alzghoul L, Al-Eitan LN, Aladawi M, Odeh M, Abu Hantash O. The Association Between Serum Vitamin D3 Levels and Autism Among Jordanian Boys. Journal of Autism and Developmental Disorders. 2020;50(9):3149-54.
129. Shamsedine L, Mailhac A, Badaoui A, El Hakim R, Kibbi R, Oueidat H, et al. Breastfeeding association with autism spectrum disorders: A case-control study from Lebanon. Research in Autism Spectrum Disorders. 2020;78.
130. Hussien KA, Roomi AB, Oudah SK. Evaluation of lead, copper and zinc levels for autistic children in thi-qar Province/Iraq. International Journal of Research in Pharmaceutical Sciences. 2020;11(3):3127-33.
131. Khalil AI, Almutairi MS, Ahmed ME. Assessing risk factors of Autism Spectrum Disorders (ASD) and Attention Deficit Hyperactivity Disorder (ADHD) among Saudi Mothers: A retrospective study. Clinical Schizophrenia and Related Psychoses. 2020;14(1).
132. Mossa ME, Ahmed AS, Khedhair KA. Evaluation of serum oxytocin hormone level in children with autism. Indian Journal of Public Health Research and Development. 2020;11(10):298-302.
133. Jenabi E, Seyedi M, Hamzehei R, Bashirian S, Rezaei M, Razjouyan K, et al. Association between assisted reproductive technology and autism spectrum disorders in iran: A case-control study. Clinical and Experimental Pediatrics. 2020;63(9):368-72.
134. Razjouyan K, Etemadi N, Miri MALI. A Study of the Prevalence of Risk Factors Associated with Autism Spectrum Disorder among Affected Individuals in Tehran. Pakistan Journal of Medical and Health Sciences. 2020;14(2):1381-4.
135. Richa S, Khoury R, Jrouhayem J, Chammay R, Kazour F, Bou Khalil R, et al. Estimating the prevalence of autism spectrum disorder in Lebanon. Encephale. 2020;46(6):414-9.
136. Virolainen S, Hussien W, Dalibalta S. Autism spectrum disorder in the United Arab Emirates: Potential environmental links. Reviews on Environmental Health. 2020;35(4):359-69.
137. Muftin NQ, Jubair S, Hadi SM. Identification of MTHFR genetic polymorphism in Iraqi autistic children. Gene Reports. 2020;18.
138. Beiranvandi F, Akouchekian M, Javadi GR, Darvish H. The association of CNTNAP2 rs2710102 and ENGRAILED-2 rs1861972 genes polymorphism and autism in Iranian population. Meta Gene. 2020;24.

139. Darvish H, Omidvar ME, Ghaedi E, Ghaedi H. Association of rs3735025 and rs9656169 variants with autism, and schizophrenia: A GWAS-replication study in an Iranian population. Meta Gene. 2020;26.
140. Kandeel WA, Meguid NA, Bjørklund G, Eid EM, Farid M, Mohamed SK, et al. Impact of Clostridium Bacteria in Children with Autism Spectrum Disorder and Their Anthropometric Measurements. Journal of Molecular Neuroscience. 2020;70(6):897-907.
141. Mobasheri L, Moossavi SZ, Esmaeili A, Mohammadoo-khorasani M, Sarab GA. Association between vitamin D receptor gene FokI and TaqI variants with autism spectrum disorder predisposition in Iranian population. Gene. 2020;723.
142. Saad K, Abdallah AEM, Abdel-Rahman AA, Al-Atram AA, Abdel-Raheem YF, Gad EF, et al. Polymorphism of interleukin-1β and interleukin-1 receptor antagonist genes in children with autism spectrum disorders. Progress in Neuro-Psychopharmacology and Biological Psychiatry. 2020;103.
143. Safari M, Noroozi R, Taheri M, Ghafouri-Fard S. The rs12826786 in HOTAIR lncRNA Is Associated with Risk of Autism Spectrum Disorder. Journal of Molecular Neuroscience. 2020;70(2):175-9.
144. Taheri M, Noroozi R, Aghaei K, Omrani MD, Ghafouri-Fard S. The rs594445 in MOCOS gene is associated with risk of autism spectrum disorder. Metabolic Brain Disease. 2020;35(3):497-501.
145. Al-Bazzaz A, A-Dahir K, Almashhadani A, Al-Ani I. Estimation of fasting serum levels of glucose, zinc, copper, zinc /copper ratio and their relation to the measured lipid profile in autistic patients and non-autistic controls in Jordan. Biomedical and Pharmacology Journal. 2020;13(1):481-8.
146. Chehbani F, Gallello G, Brahim T, Ouanes S, Douki W, Gaddour N, et al. The status of chemical elements in the blood plasma of children with autism spectrum disorder in Tunisia: a case-control study. Environmental Science and Pollution Research. 2020;27(28):35738-49.
147. Gerges P, Bitar T, Hawat M, Alameddine A, Soufia M, Andres CR, et al. Risk and protective factors in autism spectrum disorders: A case control study in the lebanese population. International Journal of Environmental Research and Public Health. 2020;17(17):1-8.
148. Soliman ES, Almutairi A, Almutairi F, Alswailem H, Salem R, Albati W. Autistic traits among Saudi university students; relations with emotional intelligence and alexithymia. European Psychiatry. 2020;63:S636-S7.

149. Al-Mamari W, Idris AB, Al-Zadjali AA, Jalees S, Murthi S, Al-Jabri M, et al. Parental age and the risk of autism spectrum disorder in Oman a case-control study. Sultan Qaboos University Medical Journal. 2021;21(3):465-71.
150. Alomar RS, Oommen A, Aljofi HE. Vitamin D deficient diet and autism. Kuwait Medical Journal. 2021;53(2):179-83.
151. Al-Ali BA, Ali AA, Mahdi SS, Kadhim ZHM, Hashim ZA. Determination of environmental risk factors of Autism in Kerbala city / Iraq 2020. Latin American Journal of Pharmacy. 2021;40:308-16.
152. Al-Sarraj Y, Al-Dous E, Taha RZ, Ahram D, Alshaban F, Tolfat M, et al. Family-based genome-wide association study of autism spectrum disorder in middle eastern families. Genes. 2021;12(5).
153. Hegazy NN, Ragab SM, Hofey WZE. Environmental risk factors associated with children autism spectrum disorders in, menoufia governorate. Egyptian Journal of Hospital Medicine. 2021;82(3):461-70.
154. Mondal A, Mukherjee S, Dar W, Singh S, Pati S. Role of glucose 6-phosphate dehydrogenase (G6PD) deficiency and its association to Autism Spectrum Disorders. Biochimica et Biophysica Acta - Molecular Basis of Disease. 2021;1867(10).
155. Nawaz FA, Sultan MA. Low Birth Weight Prevalence in Children Diagnosed with Neurodevelopmental Disorders in Dubai. Global Pediatric Health. 2021;8.
156. Al Malki JS, Hussien NA, Al Malki F. Maternal toxoplasmosis and the risk of childhood autism: serological and molecular small-scale studies. BMC Pediatrics. 2021;21(1).
157. Meguid NA, Nashaat NH, Elsaeid A, Peana M, Elnahry A, Bjørklund G. Awareness and risk factors of autism spectrum disorder in an Egyptian population. Research in Autism Spectrum Disorders. 2021;84.
158. Rahmani Z, Fayyazi Bordbar MR, Dibaj M, Alimardani M, Moghbeli M. Genetic and molecular biology of autism spectrum disorder among Middle East population: a review. Human Genomics. 2021;15(1).
159. Nakhla MAY, Zaky EA, Nabeh ES, Abd El Aziz AW. Assessment of 25 Hydroxy Cholecalceferol Level in Autistic Children; is there a role for it in Treatment of Autism Spectrum Disorder? QJM. 2021;114.
160. Shehata M, Salah E, Youssef MM, Shady MMA, El-Alameey I, Ashaat E, et al. Comparing levels of urinary phthalate metabolites in egyptian children with autism spectrum

disorders and healthy control children: Referring to sources of phthalate exposure. Open Access Macedonian Journal of Medical Sciences. 2021;9:1640-6.
161. Zaky EA, Shaaban HAM, Dawoud MOA, Madbouly KSEF, Deifalla SM. Neurodevelopmental Outcomes after Neonatal Mechanical Ventilation. QJM. 2021;114.
162. Alotaibi AM, Craig KA, Alshareef TM, AlQathmi ES, Aman SM, Aldhalaan HM, et al. Sociodemographic, clinical characteristics, and service utilization of young children diagnosed with autism spectrum disorder at a research center in Saudi Arabia. Saudi Medical Journal. 2021;42(8):878-85.
163. Sadek AA, Osman AA, Samy MN. Clinical and laboratory characteristics of children with autism spectrum disorder at sohag university hospital. Egyptian Journal of Hospital Medicine. 2021;84(1):2249-55.
164. Alkhalidy H, Abushaikha A, Alnaser K, Obeidat MD, Al-Shami I. Nutritional Status of Pre-school Children and Determinant Factors of Autism: A Case-Control Study. Frontiers in Nutrition. 2021;8.
165. Alrahili N, Almarshad NA, Alturki RY, Alothaim JS, Altameem RM, Alghufaili MA, et al. The Association Between Screen Time Exposure and Autism Spectrum Disorder-Like Symptoms in Children. Cureus. 2021;13(10):e18787.
166. Zahra SS, Mahmoud RA, Lateef RKA, Abdel Raouf BM. Socio-economic Status in Egyptian Patients with Autism Spectrum Disorder. Does it affect Autism Severity? Egyptian Journal of Hospital Medicine. 2022;89(2):6532-40.
167. Aloufi HR, Alotaibi FF, Zamzami LZ, Almadani RM. Breastfeeding and Its Relation with Autism Spectrum Disorder in Children. Egyptian Journal of Hospital Medicine. 2022;87(1):1092-6.
168. Arafa A, Mahmoud O, Salah H, Abdelmonem AA, Senosy S. Maternal and neonatal risk factors for autism spectrum disorder: A case-control study from Egypt. PLoS ONE. 2022;17(6).
169. Gholamalizadeh M, Mirsadeghi NA, Rastgoo S, Abbas Torki S, Bourbour F, Kalantari N, et al. The association of body mass index and dietary fat intake with autism in children: a case-control study. Nutrition and Food Science. 2022.
170. Kheirouri S, Farazkhah T. Contribution of Excessive Gestational Weight Gain to the Increased Risk of Autism Spectrum Disorder Occurrence in Offspring. Journal of Indian Association for Child and Adolescent Mental Health. 2022;18(1):73-81.
171. Sadiq SF, Mohsenal-Waaly AB. Toxoplasmosis is a risk factor in autism disease in Al-Diwaniyah governorate, Iraq. NeuroQuantology. 2022;20(10):9594-8.

172. Slama S, Bahia W, Soltani I, Gaddour N, Ferchichi S. Risk factors in autism spectrum disorder: A Tunisian case-control study. Saudi Journal of Biological Sciences. 2022;29(4):2749-55.

173. Akbari M, Badrlou E, Eslami S, Hussen BM, Taheri M, Neishabouri SM, et al. Association between angiotensin I converting enzyme gene polymorphisms and risk of autism in Iranian population. Human Gene. 2022;33.

174. Lord JR, Mashayekhi F, Salehi Z. How matrix metalloproteinase (MMP)-9 (rs3918242) polymorphism affects MMP-9 serum concentration and associates with autism spectrum disorders: A case-control study in Iranian population. Development and Psychopathology. 2022;34(3):882-8.

175. Hamed RMR, Ayoub MI, Abdel Samie M, Hamam NN. Anti-ganglioside M1 autoantibodies in Egyptian children with autism: a cross-sectional comparative study. Middle East Current Psychiatry. 2022;29(1).

176. Hassan MH, Shehata GA, Ahmed AE, El-Sawy SA, Tohamy AM, Sakhr HM, et al. Vitamin D3 status and polymorphisms of vitamin D receptor genes among cohort of Egyptian children with autism. Gazzetta Medica Italiana Archivio per le Scienze Mediche. 2022;181(11):824-32.

177. Raouf HA, Kholoussi N, Kholoussi S, Abo-Shanab AM, Ashaat EA, Ashaat NA, et al. Association of immune abnormalities with symptom severity in Egyptian autistic children. Egyptian Pharmaceutical Journal. 2022;21(2):242-8.

178. Rezaei M, Rezaei A, Esmaeili A, Nakhaee S, Azadi NA, Mansouri B. A case-control study on the relationship between urine trace element levels and autism spectrum disorder among Iranian children. Environmental science and pollution research international. 2022;29(38):57287-95.

179. Al-Ali Z, Yasseen AA, Al-Dujailli A, Al-Karaqully AJ, McAllister KA, Jumaah AS. The oxytocin receptor gene polymorphism rs2268491 and serum oxytocin alterations are indicative of autism spectrum disorder: A case-control paediatric study in Iraq with personalized medicine implications. PLoS ONE. 2022;17(3).

180. Al-Dakroury WA, Alnemary FM, Alnemary F. Autism in the Kingdom of Saudi Arabia: Current Situation and Future Perspectives for Services and Research. Perspectives of the ASHA Special Interest Groups. 2022;7:2104-9.

181. Aljumaili OI, El-Waseif AA, AbdulJabbar Suleiman A, El-Dein A. Ewais E. Assessment of hair aluminium, cobalt, and mercury in a specimen of autistic Iraqi patients:

Environmental risk factors of heavy metals in autism. Materials Today: Proceedings. 2022;57:350-3.
182. Alenezi S, Alnemary F, Alamri A, Albakr D, Abualkhair L. Psychotropic Medications Use among Children with Autism in Saudi Arabia. Children. 2022;9(7).
183. Dehiol RK, Dawood LJ, Alrubaee RJ. Autism spectrum disorders and electronic screen devices exposure in Al-Nasiriya city 2019-2020. Current Pediatric Research. 2022;26(4):1308-16.
184. Hajj A, Hallit S, El-Khatib R, Haidar SA, Hajj Moussa Lteif F, Hajj L, et al. Pre-, Peri-, and Neonatal Factors Associated with Autism Spectrum Disorder: Results of a Lebanese Case-control Study. Innovations in Clinical Neuroscience. 2022;19(7):22-7.
185. Aldera H, Hilabi A, Elzahrani MR, Alhamadh MS, Alqirnas MQ, Alkahtani R, et al. Do Parental Comorbidities Affect the Severity of Autism Spectrum Disorder? Cureus. 2022;14(12):e32702.
186. Sefrioui MR, Elidrissi I, El Othmani IS, Derfoufi S, Said AAH, Benmoussa A, et al. Profile of autism spectrum disorders in Morocco: cross-sectional retrospective study of parents of children with autism. Medicine and Pharmacy Reports. 2023;96(1):71-8.
187. Alakhzami M, Huang A. Individuals with Autism Spectrum Disorders and Developmental Disorders in Oman: An Overview of Current Status. Journal of Autism and Developmental Disorders. 2023;53(2):825-33.
188. Alamoudi RA, Al-Jabri BA, Alsulami MA, Sabbagh HJ. Prenatal maternal stress and the severity of autism spectrum disorder: A cross-sectional study. Developmental Psychobiology. 2023;65(2).
189. Al-Awadi BQH, Al-Aadhami MAWS, Baqer NN. Chromosomal aberration detection in Iraqi children with autism. Human Gene. 2023;38.
190. Aljumaili OI, El-Dein A. Ewais E, El-Waseif AA, AbdulJabbar Suleiman A. Determination of hair lead, iron, and cadmium in a sample of autistic Iraqi children: Environmental risk factors of heavy metals in autism. Materials Today: Proceedings. 2023;80:2712-5.
191. AlQahtani O. Autism spectrum disorder: Where does the Gulf Region stand? An overview of ASD in the Arab Gulf Region: The UAE as a regional model. Research in Autism Spectrum Disorders. 2023;108.
192. Alshaban FA, Aldosari M, Ghazal I, Al-Shammari H, ElHag S, Thompson IR, et al. Consanguinity as a Risk Factor for Autism. Journal of Autism and Developmental Disorders. 2023.

193. Heidari A, Motamed M, Rahimi Forushani A, Alaghband-Rad J. The Association Between Autism Spectrum Disorder and Attention Deficit Hyperactivity Disorder Symptoms in Medical Students. Journal of Nervous and Mental Disease. 2023;211(6):453-9.
194. Jenabi E, Ayubi E, Bashirian S, Rezaei M, Seyedi M, Razjouyan K, et al. Autism Spectrum Disorders Registry in Hamadan, Iran: A Study Protocol. Iranian Journal of Psychiatry and Behavioral Sciences. 2023;17(3).
195. Jenabi E, Seyedi M, Bashirian S, Khazaei S. Is Breastfeeding Duration Associated with Risk of Developing ASD? Current Psychiatry Research and Reviews. 2023;19(1):89-94.
196. Kassab RB, Osman A, Tarabaih A. ASSESSMENT OF CARIES PREVALENCE AND RELATED RISK FACTORS AMONG A GROUP OF LEBANESE CHILDREN WITH AUTISM SPECTRUM DISORDER: A CASE-CONTROL STUDY. International Arab Journal of Dentistry. 2023;14(2):13-21.
197. Meguid NA, Nashaat NH, Ghannoum H, Hashem HS, Hussein G, El-Saied A. Prevalence of autism spectrum disorder among children referred to special needs clinic in Giza. Egyptian Journal of Otolaryngology. 2023;39(1).
198. Nasir A, Masri A, Nasir L. Pediatricians' perspectives on childhood behavioral and mental health problems in Jordan. Middle East Current Psychiatry. 2023;30(1).
199. Saleh A, El-Kady H, Masoud M, Mohammed EA. Hair and Blood Levels of Aluminum, Cadmium, and Lead in Children with Autism from Egypt: Can Toxic Heavy Metals Increase the Risk of Autism? Iranian Journal of Toxicology. 2023;17(4):17-24.
200. Adamjee Burhani H, Hussain S, Lakshmanan J, Albanna A. Perinatal Risk Factors in Children Diagnosed with Autism Spectrum Disorder in Dubai: A Case-Control Study. Dubai Medical Journal. 2023;6(3):155-62.
201. Al-Bishri WM. Glucose transporter 1 deficiency, AMP-activated protein kinase activation and immune dysregulation in autism spectrum disorder: Novel biomarker sources for clinical diagnosis. Saudi Journal of Biological Sciences. 2023;30(12).
202. Hassan ZR, Zekry KM, Heikal EA, Ibrahim HF, Khirala SK, Abd El-Hamid SM, et al. Toxoplasmosis and cytomegalovirus infection and their role in Egyptian autistic children. Parasitology Research. 2023;122(5):1177-87.
203. Sadeghi S, Pouretemad HR, Badv RS, Brand S. Associations between Symptom Severity of Autism Spectrum Disorder and Screen Time among Toddlers Aged 16 to 36 Months. Behavioral Sciences. 2023;13(3).
204. Ebrahimi Meimand S, Amiri Z, Shobeiri P, Malekpour MR, Saeedi Moghaddam S, Ghanbari A, et al. Burden of autism spectrum disorders in North Africa and Middle East from

1990 to 2019: A systematic analysis for the Global Burden of Disease Study 2019. Brain and Behavior. 2023;13(7).
205. Abdi M, Aliyev E, Trost B, Kohailan M, Aamer W, Syed N, et al. Genomic architecture of autism spectrum disorder in Qatar: The BARAKA-Qatar Study. Genome Medicine. 2023;15(1).
206. Moravej H, Inaloo S, Nahid S, Mazloumi S, Nemati H, Moosavian T, et al. Inborn Errors of Metabolism Associated With Autism Among Children: A Multicenter Study from Iran. Indian Pediatrics. 2023;60(3):193-6.
207. Boujelben I, Chaabane M, Ben ayed I, Ben Touhemi D, Gharbi N, Guirat M, et al. Familial Autism Spectrum Disorder : A clinical study from South Tunisia. European Psychiatry. 2023;66:S390-S1.
208. Ghahari N, Yousefian F, Najafi E. Prenatal exposure to ambient air pollution and autism spectrum disorders: Results from a family-based case-control study. JCPP Adv. 2023;3(1):e12129.
209. Zebbiche Y, Stambouli B, Kaddour-Benkada I, Amziane A, Chebli AI, Achouri MY, et al. Trace element levels and autism spectrum disorder in a sample of Algerian children: A case-control study investigation. Research in Autism Spectrum Disorders. 2024;110.
